# Blunted maturation of inhibitory control circuits in the NAC-shell underlies genetic vulnerability to early-life obesity and impulsivity

**DOI:** 10.64898/2026.04.28.26351915

**Authors:** Corrado Sandini, Farnaz Delavari, Natacha Reich, Silas Forrer, Andrea Imparato, Nada Kojovic, Caren Latreche, Luigi F Saccaro, Valeria Parlatini, Samuele Cortese, Camille Piguet, Maude Schneider, Stephan Eliez, Dimitri Van De Ville

## Abstract

Obesity and behavioral impulsivity are highly heritable traits that share overlapping genetic risk and often co-occur in childhood. This study investigates whether maturation of the nucleus accumbens shell (NAC-shell) inhibitory-control circuit mediates shared vulnerability to both traits. Using state-of-the-art dynamic fMRI, we mapped NAC-shell development across 460 longitudinal assessments from childhood to adulthood, in 136 healthy controls and 126 individuals with 22q11.2 deletion syndrome (22q11DS), a genetic model conferring high risk for obesity and impulsivity. Blunted NAC-shell maturation predicted steeper BMI increases and persistent neurocognitive impulsivity, consistently mediating their association in both HC and 22q11DS populations. NAC-shell development explained both impulsivity/obesity genetic vulnerability linked to 22q11DS and familial correlations between affected/unaffected siblings. These findings propose NAC-shell maturation as a transdiagnostic endophenotype underlying obesity and self-regulation, bridging neurodevelopment, genetics, and behavior. They could inform future early-screening and precision preventive strategies for youth at risk of obesity and other impulsivity-related phenotypes.

## 1. Introduction

Due to a dramatic rise in prevalence, obesity is now responsible for an estimated 4 million deaths and 100 million disability-adjusted life years globally(1). This epidemiological trend is particularly pronounced among children and adolescents(2) and could contribute significantly to the lifelong burden of obesity-related diseases(3). Indeed, early deviations in body mass index (BMI) trajectories are among the strongest predictors of chronic adult obesity(4), underscoring the urgent need to better understand its developmental pathophysiology (5). Extensive work has examined environmental and societal contributors to the obesity epidemic and their striking geographic variation (2, 5–8). However, heritability studies estimate that 60-75% of obesity risk is genetically determined(9), with particularly high heritability in childhood(9). At first glance, such a strong genetic contribution seems at odds with the rapid and geographically heterogeneous rise in childhood obesity rates. However, intriguing evidence suggests that genetic predisposition may act by modulating sensitivity to environmental risk factors(10). The contribution of genetic factors may therefore increase proportionally to environmental risk(10), underscoring the importance of characterizing endogenous modulators that might mitigate the effects of increasingly obesogenic modern environments.

Emerging evidence suggests that deficits in inhibitory control may be a key mechanism underlying differential vulnerability to early-life obesity. Large-scale genetic and heritability studies have revealed strong covariation between high BMI and behavioral traits linked to poor impulse control(11–13). Polygenic risk scores designed to capture variation in impulsivity predict BMI (13) with almost as much accuracy as BMI-specific scores (14). Clinically, Attention-deficit/hyperactivity disorder (ADHD), a highly heritable neurodevelopmental disorder(15), is a well-replicated risk factor for obesity(16). Longitudinal studies show that ADHD often precedes the onset of obesity(17), with the risk disproportionately tied to the impulsivity-hyperactivity dimension of the disorder(18). Treatment of impulsivity with psychostimulants reduces the risk of obesity in individuals with ADHD (16, 17), suggesting a disorder-specific causal mechanism (19). This implies that early recognition and effective management of impulsivity could represent a viable strategy to protect genetically vulnerable youth from developing chronic obesity in adulthood(16, 19, 20). Nevertheless, the mechanisms connecting childhood impulsivity to later obesity remain poorly understood (21). Clarifying them could improve identification of at-risk youth and guide targeted prevention strategies.

Animal research offers promising insights into neural circuits underlying the link between impulsivity and feeding (22). The delicate balance of caloric intake and expenditure that ultimately determines body weight is regulated by a complex neural circuitry that integrates peripheral information on current nutritional status (23), with top-down information on contextual factors favoring feeding over alternative behaviors (21). The lateral hypothalamus (LH) plays a key role in integrating these inputs and promotes the execution of feeding behaviors. GABA-ergic projections from the medio-ventral shell portion of the nucleus accumbens (NAC-shell) strongly inhibit LH-mediated feeding behavior(22), potentially by blocking excitatory inputs from ventral pallidum (VP) to LH via NAC-shell-to-VP inhibitory projections(24). NAC-shell GABA-ergic outputs are tightly modulated by top-down excitatory glutamatergic inputs from prefrontal cortices (22, 25). Blocking either top-down excitatory inputs, or NAC-shell inhibitory outputs, disrupts top-down inhibitory control of feeding behavior, resulting in increased consumption of readily accessible highly caloric food, and ultimately, rapid weight gain (22). Notably, such weight gain is context-dependent, as goal-directed behavior towards non-easily accessible food is not altered (22). The NAC-shell dysconnectivity phenotype, hence, mirrors the increased food reactivity and context-dependent impulsive eating patterns that have been proposed to mediate obesity risk in ADHD(26–28). Moreover, emerging evidence suggests that a similar NAC-shell circuit may be implicated in the top-down inhibitory control of a broader set of goal-directed behaviors, mediated by the VP (24) and ventral tegmental area (VTA) (29, 30). For instance, NAC-shell-to-VTA inhibitory projections modulate the extinction of drug-seeking behaviors in animal models of addiction(31), while disrupting NAC-shell-to-VP connectivity produces a striking combination of motor hyperactivity and impulsive eating in rats(24). Together, these findings indicate that disturbances in NAC-shell connectivity can explain the link between behavioral impulsivity and obesity.

The few existing studies of this circuit in humans support this hypothesis. Reductions in NAC-shell to prefrontal connectivity have been reported in adults with binge eating disorder(32), while case studies suggested that deep brain stimulation of NAC-shell is effective in treating both treatment-resistant binge-eating(32) and drug addiction(33). Together, these findings implicate NAC-shell dysconnectivity as a neural pathway linking impulsivity and obesity, but this hypothesis has not yet been directly tested in humans. Moreover, because existing work focuses on adults, the developmental trajectories of NAC-shell function remain unknown. It is unclear whether atypical NAC-shell maturation contributes to childhood impulsivity, common in ADHD (34), and to progressive BMI deviations, which often emerge during early adolescence(35) and precede chronic adult obesity(36). Finally, it remains unknown whether NAC-shell dysfunction mediates shared genetic vulnerability to impulsivity and obesity (10, 13). Demonstrating such an endophenotypic mediating role would position NAC-shell maturation as a promising therapeutic target for mitigating the impact of genetic obesity risk.

To address these gaps, we first identified the NAC-shell inhibitory circuit in human resting-state fMRI using a recently developed dynamic functional connectivity framework(37), uniquely suited for this purpose. We validated this circuit in two independent adult samples and characterized its normative developmental trajectory in a large longitudinal cohort of healthy controls (HCs) followed from childhood to adulthood (37). We compared this trajectory with that of individuals with 22q11.2 deletion syndrome (22q11DS)(38), a genetic disorder conferring high genetic risk for both ADHD(39) and obesity(40). We recently reported that genetic vulnerability to adult obesity in 22q11DS is largely mediated by childhood impulsivity (41). Here, we tested whether atypical NAC-shell maturation accounts for both behavioral impulsivity and accelerated BMI growth, and whether it mediates their association. We found that blunted NAC-shell development indeed predicted higher impulsivity and steeper BMI trajectories, fully mediating their association in 22q11DS. We then examined inter-individual variability in NAC-shell maturation observed among healthy controls, revealing that this circuit robustly explains population-wide co-variation of self-regulation deficits and obesity risk, beyond 22q11DS. Finally, we assessed whether NAC-shell maturation reflects shared genetic liability for childhood obesity and impulsivity. Our results indicate that this circuit operates as a transdiagnostic endophenotype mediating both the specific genetic risk associated with 22q11DS and additional heritable influences shared among affected and unaffected siblings. Together, these findings help clarify the neurodevelopmental mechanisms linking genetic vulnerability to impulsivity and obesity and provide a mechanistic basis for early targeted interventions.

## 2. Results

### 2.1 A NAC-shell inhibitory control circuit identified in healthy adults

We first investigated whether the NAC-shell inhibitory control circuit could be reliably identified from resting-state fMRI data across independent healthy adult discovery and replication samples (HC-ds and HC-rs), using a recently developed micro-coactivation patterns (μ*CAPs*) approach uniquely suited for this purpose. Indeed, unlike previous functional dissection methods of striatal circuits (42), the μ*CAPs* approach incorporates both positive and negative coactivation patterns rather than disregarding co-deactivations between brain regions (37). This feature is particularly advantageous for studying the NAC-shell’s role as a top-down inhibitory relay, which involves antagonistic coordination between brain regions. Animal data further suggest that distinct NAC circuits often overlap anatomically, forming interwoven information channels rather than clearly segregated structures (31). μ*CAPs* are uniquely suited for differentiating anatomically overlapping yet functionally distinct information flow channels, as they do not impose discrete anatomical or temporal boundaries among identified circuits (37).

Applied to the NAC, the approach revealed six μ*CAPS* reminiscent of neuronal circuits characterized by distinct activation gradients within the seed region, each associated with a unique whole-brain coactivation pattern (see Supplementary Figure-1). One of the μ*CAPs* emerged as a strong candidate for the NAC-shell inhibitory control circuit; i.e., its within-seed activation gradient was centered in the ventral–posterior–inferior portion of the NAC, consistent with previous histological(43) and structural connectivity-based segmentations of the NAC-shell (32, 44). The corresponding whole-brain pattern closely matched NAC-shell circuitry described in animal models (22, 31) (Figure-1-Panel-A). Specifically, this candidate for the NAC-shell circuit showed strong negative coactivation with the Ventral Tegmental Area (VTA), consistent with inhibitory projections from the NAC-shell to the VTA (22, 31). Positive coactivation was observed in the anterior hypothalamus, whereas the posterior–lateral hypothalamus (PLH), involved in human feeding control (45–48), displayed strong negative coactivation, mirroring the PLH-inhibitory effects of GABA-ergic NAC-shell projections in animal studies (22). Additional negative coactivations were observed in limbic regions involved in emotional processing, including the insular cortex and amygdala, again consistent with inhibitory connections from the NAC-shell to emotion-related limbic areas(49). Finally, strong positive coactivation was observed across distributed frontoparietal regions, consistent with the NAC-shell’s role in mediating prefrontal inhibitory control of VTA-mediated goal-directed (29, 50) and PLH-mediated feeding behavior (22). Results from μ*CAPs*-based dissections were highly consistent across both discovery and replication samples (see Supplementary Figure-2/3).

**Figure 1.**
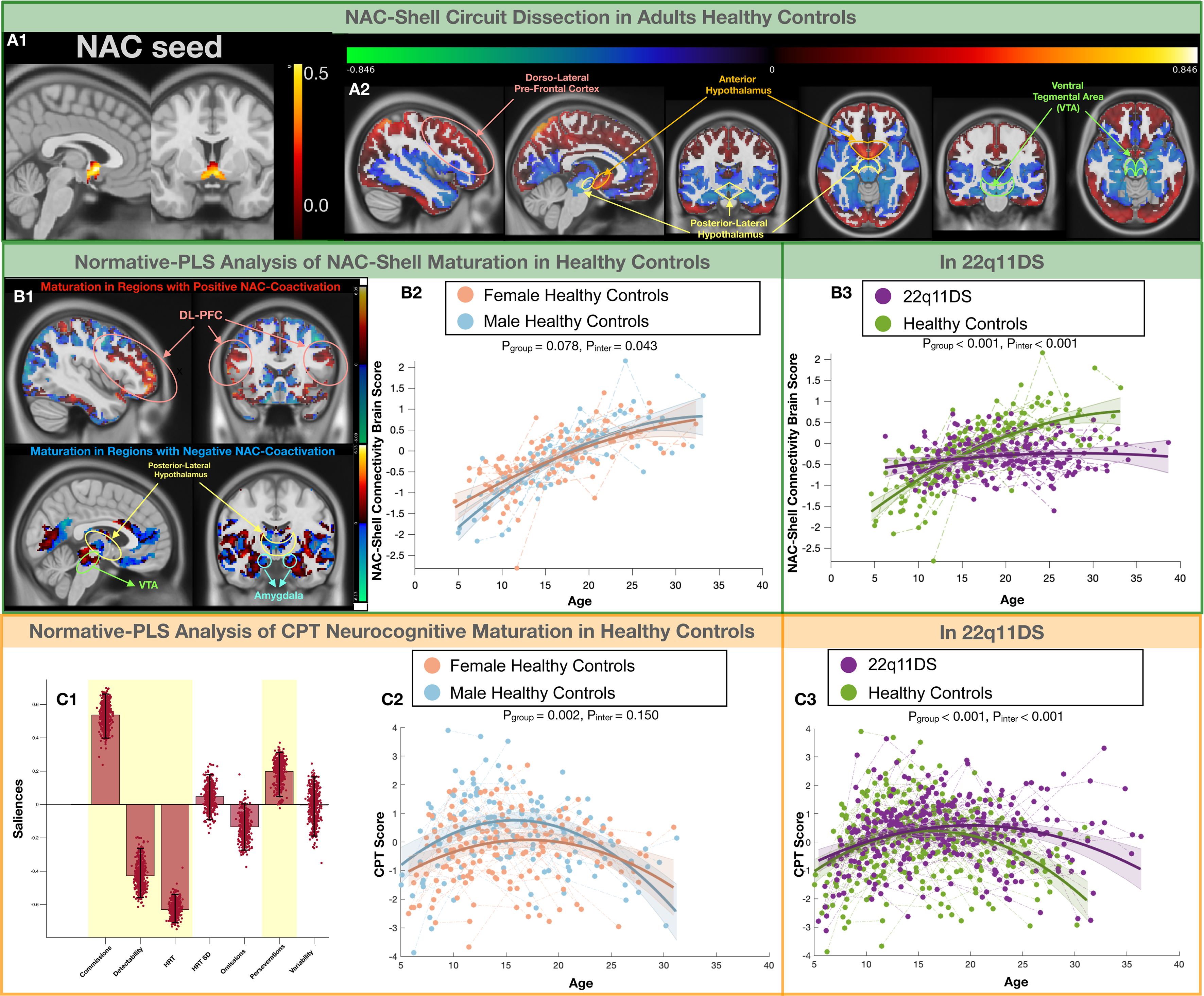
μCaps Functional Dissection of the NAC-Shell Circuit in Adult Healthy Controls and Normative PLS Analysis of NAC-Shell and Neurocognitive Maturation Panel A: NAC-Shell functional circuit dissection in adult healthy controls. **Panel A1:** Activation pattern within the NAC seed, showing which portion of the NAC contributed to the overall seed activation pattern across selected frames. The within-seed activation gradient is centered in the ventral-anterior portion of the NAC, consistent with histological and structural connectivity-based segmentations of the NAC shell. **Panel A2:** Whole-brain co-activation patterns representing average fMRI activity during frames assigned to the corresponding candidate NAC-Shell circuit. Red shading indicates positive co-activations with the ventral/anterior Shell portion of the NAC, while blue shading indicates negative co-activations, meaning that those brain regions exhibited lower-than-average activity during frames when the NAC-Shell seed was most active. The anatomical pattern revealed strong positive co-activations in higher-order fronto-parietal cortices, particularly in the dorsolateral prefrontal cortex (highlighted in pink), together with negative co-activations in the ventral tegmental area (VTA; highlighted in green) and posterior-lateral hypothalamus (highlighted in yellow). This candidate NAC-Shell circuit aligns with the known functional role of the NAC shell in mediating top-down prefrontal inhibitory control over both VTA-driven goal-directed behavior and feeding behavior mediated by the posterior-lateral hypothalamus. **Panel B:** Normative PLS analysis of NAC-Shell circuit maturation in healthy controls and comparison with the 22q11DS sample. **Panel B1:** Multivariate brain pattern describing age-related maturation of the NAC-Shell circuit in healthy controls. Red shading reflects a positive effect of age, while blue shading reflects a negative effect. The upper brain maps show regions with overall positive co-activation patterns with the NAC shell in adults; a positive effect of age indicates progressive strengthening and maturation of these positive co-activations, as observed in the dorsolateral prefrontal cortex (highlighted in pink). The lower maps depict regions with significant negative co-activation patterns with the NAC shell in adults. Here, blue shading indicates strengthening and maturation of these negative co-activations, as observed in the posterior-lateral hypothalamus (highlighted in yellow), posterior portions of the VTA (highlighted in green), and the amygdala (highlighted in light blue). **Panel B2:** Developmental trajectories of NAC-Shell Maturation Scores (NAC-Shell-MS) derived from normative PLS analysis, compared across male and female healthy controls. **Panel B3:** Developmental trajectories of NAC-Shell-MS compared between healthy controls (HCs) and individuals with 22q11DS. **Panel C:** Normative PLS analysis of CPT neurocognitive maturation in healthy controls. **Panel C1:** Multivariate CPT pattern describing the contribution of each variable to the corresponding neurocognitive dimension. Variables highlighted in yellow contribute significantly to the pattern. The direction of the bar reflects the direction of association (upward for positive and downward for negative). The pattern is characterized by a negative contribution of Hit Reaction Time (indicating faster response propensity), positive contributions of Commission and Perseveration Errors, and a negative contribution of Detectability, indicating difficulty in discriminating target stimuli from distractors, collectively suggestive of a more impulsive response style. **Panel C2:** Developmental trajectories of CPT Impulsivity Maturation Scores (CPT-Impulsivity-MS) derived from normative PLS analysis, compared across male and female healthy controls. **Panel C3:** Developmental trajectories of CPT-Impulsivity-MS compared between HCs and individuals with 22q11DS.

### 2.2 Normative NAC-shell circuit maturation trajectory in healthy controls

We next characterized the normative maturational trajectory of the NAC-shell-associated μCAP in a longitudinal sample of 136 healthy controls (HCs), aged 6–34 years, spanning 227 assessments. For each scan, frames showing significant NAC activation were matched to the closest adult μCAP, and average NAC-shell coactivation maps were computed by averaging all matched frames. We applied a longitudinal partial least squares (PLS) correlation analysis to characterize multivariate patterns of age-related maturation of the NAC-shell circuit.

The PLS analysis revealed a single significant component (R=0.6, p<0.001 Figure-1-Panel-B) capturing age-related NAC-shell maturation in HCs. Brain loadings showed progressive strengthening of positive coactivation between the NAC-shell and prefrontal cortices, combined with strengthening of negative coactivation with the posterior-lateral hypothalamus, amygdala, and portions of the VTA (Figure-1-Panel-B1). To quantify individual variability, we computed a NAC-shell Maturation Score (NAC-shell-MS) for each assessment by multiplying PLS-derived loadings with each subject’s NAC-shell coactivation pattern. This single index reflected the degree to which each assessment aligned with the normative trajectory. Modeling NAC-shell-MS as a function of age and gender showed a prolonged developmental course continuing into early adulthood (Figure-1-Panel-B2), consistent with extended maturation of top-down inhibitory control (51). Males exhibited a slight but significant delay relative to females (P-Group-Effect=0.078, P-Age-Interaction=0.043 Figure-1-Panel-B2), mirroring the higher prevalence of impulsivity symptoms in boys during childhood (52).

### 2.3 NAC-Shell circuit maturation in a 22q11DS genetic high-risk sample

To characterize the functional correlates of circuit maturation, we compared the normative NAC-Shell trajectory defined in HCs with that observed in a longitudinal genetic high-risk sample with 22q11.2 Deletion Syndrome (22q11DS; 126 individuals with 233 longitudinal assessments). Using brain loadings derived from the normative PLS analysis, we computed a NAC-shell-MS for each longitudinal assessment in the 22q11DS group, applying the same matrix-multiplication approach used for the HCs. We then compared NAC-shell-MS trajectories between the two groups. For both groups, each individual’s mean NAC-shell-MS was also compared against the normative age- and sex-adjusted trajectory derived from the HC sample. Differences in the proportion of individuals classified as having atypically low NAC-shell-MS across samples were used to estimate the impact of 22q11DS-related genetic vulnerability on circuit maturation. Moreover, by comparing typical versus atypical NAC-shell subgroups within both cohorts, we evaluated transdiagnostic effects on obesity and impulsivity-related phenotypes.

Compared with HCs, the 22q11DS group showed markedly lower NAC-shell-MS values from childhood, with divergence increasing over time due to a blunted developmental slope (P-Group-Effect<0.001, P-Age-Interaction<0.001, Figure-1-Panel-B3). Consequently, a larger proportion of 22q11DS participants exhibited atypical below-average NAC-shell-MS compared with HCs (Typical/High-22q11=40,M/F=23/17, Atypical/Low-22q11=86,M/F=42/44, Typical/High-HC=69,M/F=31/38, Atypical/Low-HC=67,M/F=31/36, Chi^2^=9.71, p=0.002). Modeling NAC-shell-MS across subgroups revealed a transdiagnostic gradient of maturation scores: Typical-HC>Typical-22q11DS>Atypical-HC>Atypical-22q11DS (Figure-3-Panel-A), indicating that 22q11DS increases both the prevalence and severity of NAC-shell maturation deficits. These subgroupings were subsequently used to examine behavioral and metabolic correlates of NAC-shell maturation in both populations.

### 2.3 Transdiagnostic effects of NAC-shell circuit maturation on BMI trajectories

BMI trajectories diverged significantly across both HC and 22q11DS NAC-shell subgroups. Progressive BMI increases appeared in Atypical-NAC-shell-HC relative to Typical-NAC-shell-HC (P-Group-Effect=0.02, P-Age-Interaction=0.01, Figure-3-Panel-C2) and in Atypical-NAC-shell-22q11DS relative to Typical-NAC-shell-22q11DS (P-Group-Effect=0.002, P-Age-Interaction=0.001, Figure-3-Panel-C3.). In both cohorts, divergence peaked during late childhood and early adolescence, closely matching the anticipation of adiposity rebound, previously identified as a strong predictor of adult obesity (4, 35). BMI trajectories continued to separate across adolescence, resulting in higher adult BMI scores in the Atypical-than in the Typical-NAC-shell subgroup.

Four group analysis revealed a transdiagnostic gradient that inversely mirrored NAC-shell-MS gradient, with progressively greater BMI increases in the order: Atypical-NAC-shell-22q11DS>Atypical-NAC-shell-HC>Typical-NAC-shell-22q11DS>Typical-NAC-shell-HC (P-Group-Effect<0.001, P-Age-Interaction<0.001, Figure-3-Panel-C1). As a complementary step, we derived a normative Longitudinal-BMI-Trajectory Score by comparing each participant’s BMI rate-of-change with the gender-matched normative trajectory. Positive values indicated steeper-than-average BMI growth, and negative values indicated flatter-than-average BMI growth. This Longitudinal-BMI-Trajectory Score correlated negatively with NAC-shell-Maturation Score across the entire sample (ρ=-0.35, p-value<0.001, Figure-3-Panel-C4), within 22q11DS (ρ=-0.66, p<0.001) and within HC populations (ρ=-0.33, p=0.001). These findings indicate that individual variation in NAC-shell maturation is directly, and transdiagnostically, linked to vulnerability to accelerated BMI growth, conferring greater risk for adult obesity.

### 2.4 Normative neurocognitive impulsivity trajectories and effects of 22q11DS genetic vulnerability

To examine the potential impact of NAC-shell maturation, we first characterized the normative trajectory of neurocognitive inhibitory control, assessed with the Conners Continuous Performance Test (CPT). CPT scores reflect multiple ADHD-related cognitive dimensions, including attention and inhibition, which can be separated through multivariate latent structure analysis of test items (53). To model normative developmental trajectories of these dimensions, we analyzed age and sex effects on raw CPT scores in HCs using longitudinal multivariate PLS analysis. From this model, we derived neurocognitive maturation scores for each assessment in both HC and 22q11DS samples, employing the same matrix-multiplication procedure as for the NAC-shell-MS.

The normative CPT-PLS analysis identified two significant components reflecting distinct developmental trajectories (Figure-1-Panel-C and Supplementary-Figure-4-Panel-A). The first component (R=0.64, p=0.001, Supplementary-Figure-4-Panel-A), detailed in the Supplementary Material, captured progressive improvements in attention performance reflected by reductions in omission, commission, and perseveration errors, improved detectability, and decreased Hit Reaction Time (HRT) and variability. The corresponding CPT-Attention Maturation Score (CPT-Attention-MS) increased sharply through childhood and early adolescence and leveled out in early adulthood. This score was lower in 22q11DS regardless of age, consistent with syndrome-related attentional difficulties (39, 54–56), although age-related maturation trajectories were comparable across groups (P-Group-Effect=0.005, P-Age-Interaction=0.4, R=0.64, p=0.001, Supplementary-Figure-4-Panel-A3).

The **s**econd developmental component (R=0.29, p=0.001, Figure-1-Panel-C) was strongly indicative of neurocognitive impulsivity. This component was characterized by positive loadings on commission and perseveration errors and negative loadings on detectability and HRT (Figure-1-Panel-C1), reflecting a diminished ability to inhibit reflexive responses, resulting in excessively short reaction times and reduced discrimination between targets and distractors (57). Developmentally, impulsivity followed an inverse U-shaped trajectory, peaking during adolescence and declining into adulthood. Impulsivity levels were higher in males than females (P-Group-Effect=0.002, P-Age-Interaction=0.15, Figure-1-Panel-C2), consistent with adolescent increase in behavioral impulsivity and risk-taking (58, 59), and known sex differences in impulsivity (52, 60). The CPT-Impulsivity-Maturation Score was also elevated since childhood in 22q11DS (P-Group-Effect<0.001, P-Age-Interaction<0.001), with a further failure to show the normative decline in impulsivity during early adulthood observed in HCs (Figure-1-Panel-C3).

### 2.4 Transdiagnostic effects of NAC-shell circuit maturation on neurocognitive impulsivity trajectory

Variance in NAC-shell maturation was only mildly associated with blunting of the CPT-Attention-MS trajectory in the 22q11DS Atypical-NAC-shell-MS subgroup compared with the Typical subgroup (P-Group-Effect=0.049, P-Age-Interaction=0.02, Supplementary-Figure-4-Panel-B3), and had no significant effect in HCs (P-Group-Effect=0.85, P-Age-Interaction=0.7, Supplementary-Figure-4-Panel-B2).

In contrast, NAC-shell maturation was strongly associated with CPT-Impulsivity-MS trajectories in both HC (P-Group-Effect<0.001, P-Age-Interaction<0.001, Figure-2-Panel-B2) and 22q11DS populations (P-Group-Effect=0.002, P-Age-Interaction=0.003, Figure-2-Panel-B3). A transdiagnostic gradient of CPT-Impulsivity-MS emerged that closely mirrored NAC-shell subgrouping, with the highest impulsivity levels in Atypical-NAC-shell-22q11DS>Atypical-NAC-shell-CS>Typical-NAC-shell-22q11DS>Typical-NAC-shell-HC (P-Group-Effect<0.001, P-Age-Interaction<0.001, Figure-2-Panel-B). This relationship was supported by a negative correlation between mean NAC-shell-MS and CPT-Impulsivity-MS (ρ=-0.26, p<0.001 Figure-2-Panel-B4). The influence of NAC-shell maturation on neurocognitive impulsivity was remarkably consistent across both HC and 22q11DS populations. Atypical-NAC-shell subgroups exhibited greater CPT-Impulsivity-MS during childhood and a weaker decline in impulsivity from adolescence to early adulthood compared with their Typical-NAC-shell counterparts. These findings highlight NAC-shell maturation as a shared determinant of developmental impulsivity trajectories in both HC and 22q11DS populations.

**Figure 2.**
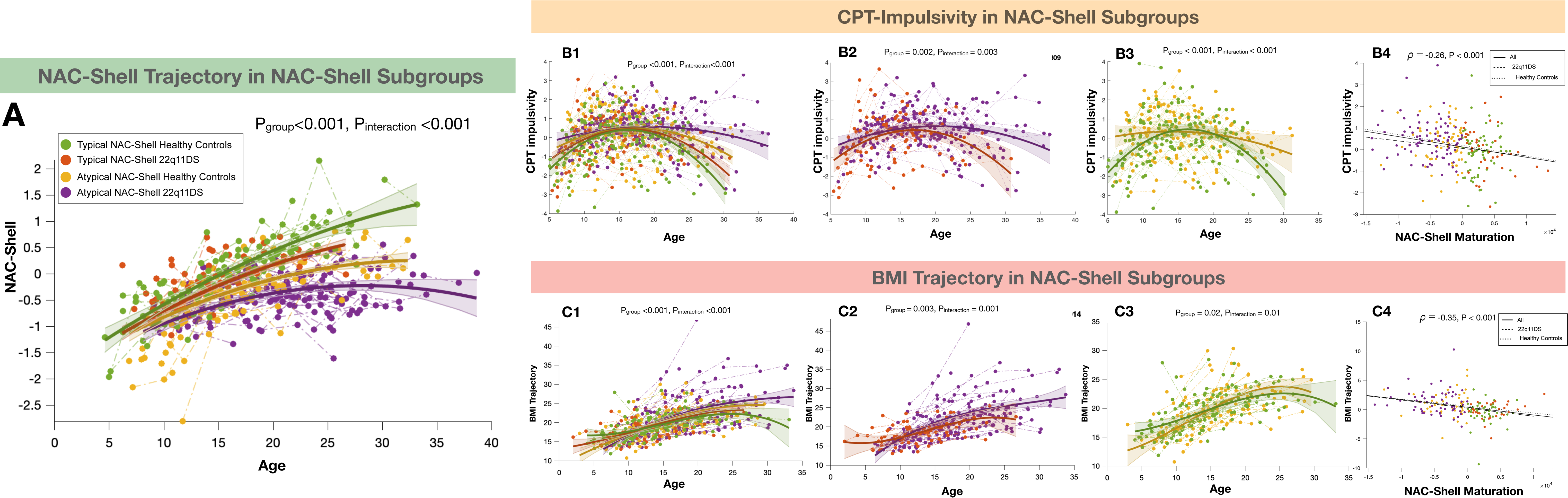
Transdiagnostic Correlates of Typical vs. Atypical NAC-Shell Maturation Across HC and 22q11DS Samples Panel A: Trajectories of NAC-Shell Maturation Scores compared across transdiagnostic NAC-Shell subgroups (Atypical-NAC-Shell-22q11DS in purple, Atypical-NAC-Shell-HC in yellow, Typical-NAC-Shell-22q11DS in orange, and Typical-NAC-Shell-HC in green). **Panel B1:** Trajectories of CPT-Impulsivity-MS across transdiagnostic NAC-Shell subgroups. **Panel B2:** Trajectories of CPT-Impulsivity-MS compared between typical and atypical NAC-Shell maturation subgroups within HCs. **Panel B3:** Trajectories of CPT-Impulsivity-MS compared between typical and atypical NAC-Shell maturation subgroups within 22q11DS. **Panel B4:** Association between average NAC-Shell Maturation Score and average CPT-Impulsivity Maturation Score. **Panel C1:** BMI trajectories compared across transdiagnostic NAC-Shell subgroups. **Panel C2:** BMI trajectories compared between typical and atypical NAC-Shell maturation subgroups within HCs. **Panel C3:** BMI trajectories compared between typical and atypical NAC-Shell maturation subgroups within 22q11DS. **Panel C4:** Association between average NAC-Shell Maturation Score and the longitudinal BMI Trajectory Score, derived by comparing each individual’s BMI rate of change to the normative, gender-matched trajectory observed in HCs.

### 2.5 NAC-shell circuit maturation links childhood impulsivity to obesity risk, mediating 22q11DS genetic risk

Having observed robust transdiagnostic associations with both impulsivity and deviant BMI trajectories, we next examined whether atypical NAC-shell maturation mediated the link between these two phenotypes. A formal mediation analysis confirmed that NAC-shell-MS significantly mediated the association between average CPT-Impulsivity-MS and longitudinal BMI-Trajectory Scores (β_a*b_ = 0.06, 95% CI = [0.02–0.10], p = 0.02 Figure-3-Path-1).

**Figure 3.**
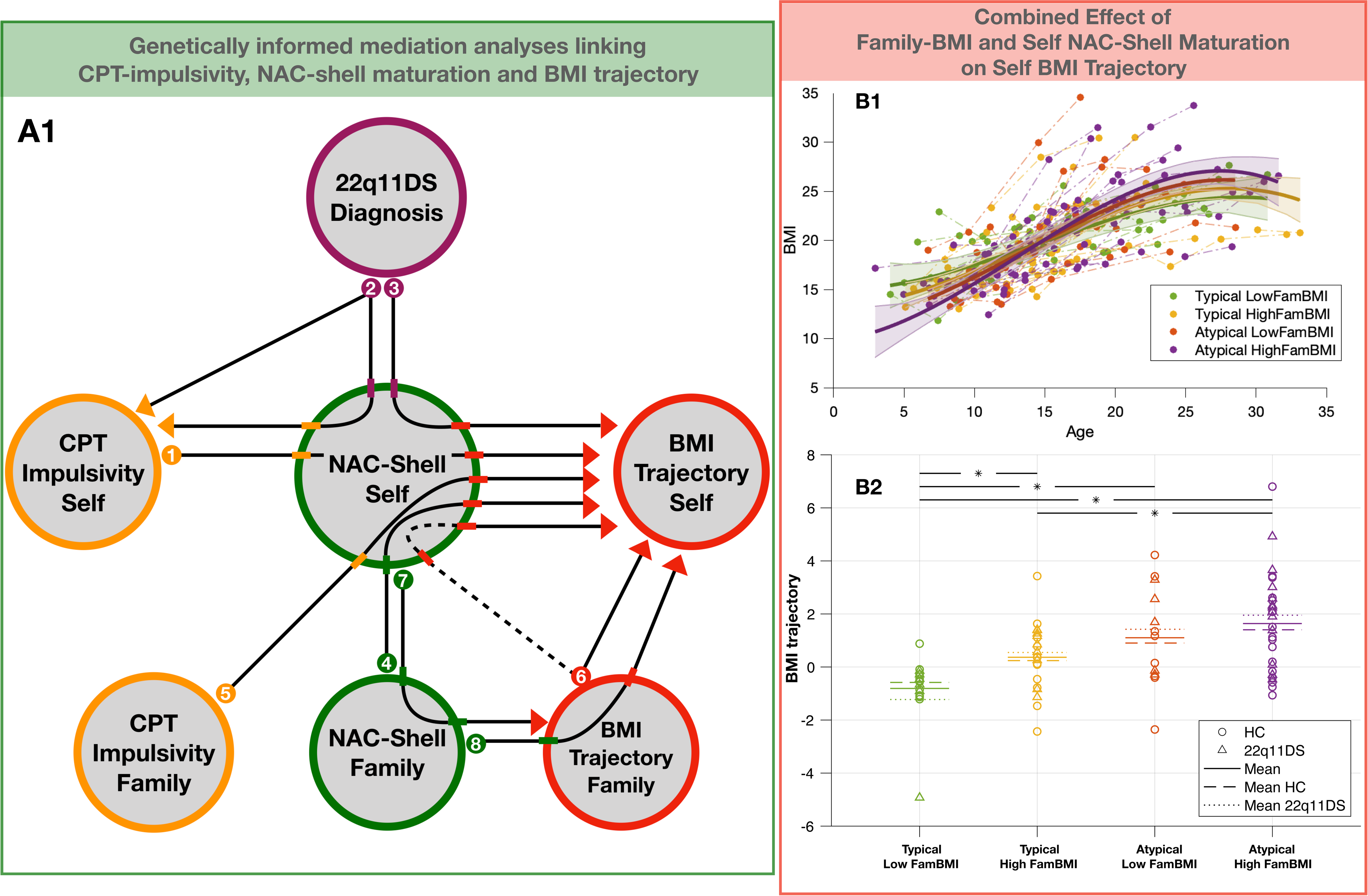
Genetically Informed Mediation Analyses Linking CPT Impulsivity, NAC-Shell Maturation, and BMI Trajectory Panel A1: Graphical summary of eight mediation analyses described in the main text. *Path 1*: NAC-Shell Maturation Score (NAC-Shell-MS) mediating the link between CPT-Impulsivity Maturation Score (CPT-Impulsivity-MS) and BMI trajectory. *Path 2*: NAC-Shell-MS mediating the effect of 22q11DS diagnosis on CPT-Impulsivity-MS. *Path 3*: NAC-Shell-MS mediating the effect of 22q11DS diagnosis on BMI trajectory. *Path 4*: NAC-Shell-MS mediating the link between Familial NAC-Shell-MS and BMI trajectory. *Path 5*: NAC-Shell-MS mediating the link between Familial CPT-Impulsivity-MS and BMI trajectory. *Path 6*: NAC-Shell-MS mediating the link between Familial BMI trajectory and individual BMI trajectory. *Path 7*: Familial NAC-Shell-MS mediating the link between NAC-Shell-MS and Familial BMI trajectory. *Path 8*: Familial BMI trajectory mediating the link between Familial NAC-Shell-MS and BMI trajectory. **Panel B:** Combined effects of familial BMI and self NAC-Shell maturation on individual BMI trajectory. **Panel B1:** BMI trajectories compared across combined Familial-BMI and Self-NAC-Shell subgroups (Typical-NAC/Low-Familial-BMI in green, Typical-NAC/High-Familial-BMI in yellow, Atypical-NAC/Low-Familial-BMI in orange, and Atypical-NAC/High-Familial-BMI in purple). **Panel B2:** Pairwise comparisons of BMI trajectory scores across combined Familial-BMI and Self-NAC-Shell subgroups.

Although these effects were evident across both groups, the 22q11DS cohort showed roughly twice the likelihood of atypical NAC-shell maturation. We therefore tested whether this atypical pattern could mediate the elevated genetic risk for impulsivity and early-life obesity associated with 22q11DS. Mediation analyses showed that vulnerability to atypical NAC-shell maturation was a significant partial mediator of the 22q11DS effect on CPT-Impulsivity-MS (β_a*b_= 0.05, 95% CI = [0.02–0.1], *p* = 0.002, Figure-3-Path-2), and a complete mediator of its effect on longitudinal BMI-Trajectory Scores (β_a*b_ = 0.05, 95% CI = [0.01–0.11], *p*= 0.008, Figure-3-Path-3). Notably, ∼30% of 22q11DS individuals with typical NAC-shell maturation displayed BMI and CPT-Impulsivity-MS trajectories indistinguishable from HC. Together, these findings suggest that genetic susceptibility to atypical NAC-shell maturation directly underlies the increased vulnerability to both neurocognitive impulsivity and early-onset obesity observed in 22q11DS.

### 2.5 NAC-shell circuit maturation as a familial endophenotype linking impulsivity and obesity risk

While NAC-shell maturation largely accounted for the effects of 22q11DS genetic vulnerability, 22q11DS-related factors alone could not explain the wide variance in NAC-shell maturation within both 22q11DS and HC groups. We therefore examined whether genetic influences outside the 22q11.2 locus contribute to NAC-shell variability and shared familial risk for obesity and impulsivity. To this end, we leveraged the family-based design of our cohort by focusing on families with both 22q11DS participants and their non-affected siblings. We correlated each participant’s NAC-shell-MS with the mean NAC-shell-MS of their siblings across 53 families (mean siblings per family = 2.4 ± 0.63). Parallel correlations were computed for BMI-Trajectory Scores (32 families, mean = 2.5 ± 0.72) and CPT-Impulsivity-MS (51 families, mean = 2.4 ± 0.63).

Significant familial correlations emerged between Self and Family NAC-shell-MS (ρ = 0.25, p = 0.004, Supplementary-Figure-5-Panel-A2), Self and Family BMI-Trajectory Scores (ρ = 0.36, p = 0.001, Supplementary-Figure-5-Panel-A3), and Self and Family CPT-Impulsivity-MS (ρ = 0.2, p = 0.03, Supplementary-Figure-5-Panel-A1). Additional cross-trait associations, between Family-BMI-Trajectory and Family-NAC-shell-MS, Family-NAC-shell-MS and Self-BMI-Trajectory, and Family-CPT-Impulsivity-MS and Self-NAC-shell-MS (see Supplementary Table-1), support shared familial influences on NAC-shell maturation, impulsivity, and BMI trajectories.

We next conducted mediation analyses (Figure-3; see Supplementary Table-2) to test whether familial effects on BMI and impulsivity operate through NAC-shell maturation. Results indicated that Self-NAC-shell-MS mediated the effects of both Family-NAC-shell-MS (β_a*b_= –0.08, 95% CI = [–0.17 to –0.02], p = 0.06; Figure-3-Path-4) and Family-CPT-Impulsivity-MS (β_a*b_= –0.08, 95% CI = [–0.01 to –0.18], p=0.002; Figure-3-Path-5) on Self-BMI-Trajectory. Only the association between Familial and Self BMI-Trajectory Scores, remained significant when accounting for Self-NAC-shell-MS (β_c’_ = 0.29, 95% CI = [0.08 to 0.49], p = 0.007; Figure-3-Path-6), although Self-NAC-shell-MS also contributed a highly significant effect on Self-BMI when accounting for Family-BMI-Trajectory (β_b_= −0.32, 95% CI = [−0.52 to −0.12], p = 0.002). The association between Self-NAC-shell-MS and Family-BMI-Trajectory was mediated by Family-NAC-Shell-MS (β_a*b_ = –0.1, 95% CI = [–1.89 to –0.02], p = 0.002; Figure-3-Path-7), while Family-BMI-Trajectory, contributed to mediate the effects of Family-NAC on Self-BMI (β_a*b_ = –0.14, 95% CI = [–0.29 to –0.03], p=0.002 Figure-3-Path-8). Overall, these findings suggest that NAC-shell maturation reflects heritable influences shared among siblings that directly contribute to familial co-occurrence of impulsivity and atypical BMI. Familial factors, however, still contributed an additional effect of BMI trajectory, suggesting the existence of additional independent mechanisms.

To assess the combined impact of familial BMI and individual NAC-shell maturation, we compared BMI trajectories across subgroups defined by Self NAC-shell maturation (Typical vs. Atypical) and Familial BMI trajectory (Low vs. High). Pronounced differences emerged (P-Group-Effect<0.0001, P-Age-Interaction<0.0001 Figure-3-Panel-B1), with the steepest BMI increases in the High-Family-BMI/Atypical-NAC-shell subgroup, followed by Low-Family-BMI/Atypical-NAC-shell, High-Family-BMI/Typical-NAC-shell, and Low-Family-BMI/Typical-NAC-shell groups. Pairwise analyses showed that typical-vs-atypical NAC-shell maturation significantly affected BMI Trajectory Scores under both Low-Family-BMI (p < 0.01, Cohen’s D = −1.219) and High-Family-BMI (p=0.02, Cohen’s D = −0.774) conditions (Figure-3-Panel-B2). In contrast, familial BMI influenced BMI trajectories only among individuals with Typical NAC-shell maturation (p<0.01, Cohen’s D = −0.947), not in Atypical subgroups (p=0.58, Cohen’s D = −0.28; Figure-3-Panel-B2).

Together, these findings indicate that Atypical NAC-shell maturation exerts a strong, independent effect on BMI trajectories, regardless of familial BMI, whereas familial BMI contributes additional risk only when NAC-shell maturation is typical.

Overall, NAC-shell maturation emerges as a shared neurodevelopmental endophenotype mediating both 22q11DS-related and broader familial genetic vulnerabilities for obesity and impulsivity.

## 3. Discussion

The rapid rise global in obesity and its marked regional variability, support environmental and societal drives of obesity risk (2, 5–8). While the role of increasingly obesogenic environments is well established (2, 5–8), growing evidence indicates that endogenous factors modulate individual vulnerability to these pressures (10). Such modulators may explain both the high heritability of obesity (9) and the amplification of genetic effects in obesogenic contexts (10). Here, we examined the maturation of the NAC-shell inhibitory-control circuit as a candidate neurodevelopmental endophenotype linking genetic obesity risk with epidemiological (61), genetic(10, 11, 13), and clinical associations (16, 17, 19, 20) between obesity and impulsivity.

Although the neurobiology of human obesity remains incompletely understood, animal data implicate a top-down inhibitory circuit connecting prefrontal cortices, the NAC-shell, and the posterior-lateral hypothalamus (PLH), which regulates feeding (22). This pathway enables prefrontal regions to suppress feeding when alternative behaviors are appropriate (22). Disrupting this circuit induces excessive eating and weight gain effects modulated by caloric content and accessibility of available food, thus paralleling impulsive eating (22). Few human studies have explored this mechanism, yet they support links between prefrontal–NAC-shell connectivity and binge eating (32) or related impulsive behaviors (33). However, characterizing NAC-shell function in humans is technically challenging because the circuit’s components are anatomically interwoven (31) and functionally antagonistic (22, 49, 50).

We addressed these challenges using the μCAPs approach (37), which allowed us to delineate a NAC-shell inhibitory circuit mirroring animal findings (22). This circuit exhibited positive co-activation with prefrontal cortices, consistent with top-down excitatory projections(25), and negative co-activation with specific posterior-lateral hypothalamic regions involved in human feeding control (45–48). This configuration was robust across independent adult samples, validating its reproducibility. We then characterized its normative developmental trajectory in HCs. Taking a developmental perspective is crucial, as vulnerability to adult obesity often arises from early deviations in BMI in late childhood or early adolescence (4). Our findings confirmed a dynamic, protracted circuit maturation, with increasing prefrontal–NAC-shell co-activation and stronger NAC-shell–PLH inhibition, consistent with gradual strengthening of top-down inhibitory control (51).

We then examined whether inter-individual differences in NAC-shell circuit maturation could serve as an endophenotype mediating genetic risk for early-life obesity. To this end, we leveraged a unique longitudinal cohort of individuals with 22q11.2 deletion syndrome (22q11DS) (38), a well-characterized genetic disorder associated with early and progressive BMI deviations(40) and elevated adult obesity risk(62). 22q11DS exerted a profound impact on NAC-shell maturation, nearly doubling the proportion of individuals exhibiting atypically low NAC-shell Maturation Scores (NAC-shell-MS) and increasing overall deficit severity. Interestingly, significant variance in NAC-shell maturation was also observed in HCs. Longitudinal analyses revealed that BMI trajectories diverged across NAC-shell subgroups in both HC and 22q11DS populations. Across both groups, atypical NAC-shell maturation predicted steeper BMI increases, whereas 22q11DS participants with typical maturation exhibited BMI trajectories similar to HCs. Mediation analyses confirmed that NAC-shell maturation fully accounted for the effect of 22q11DS on BMI growth, identifying it as an endophenotype linking genetic risk to obesity.

To clarify the mechanisms connecting circuit maturation and metabolic outcomes, we examined impulsivity. In animal models, the NAC-shell not only regulates feeding but also suppresses goal-directed behaviors through inhibitory projections to the ventral tegmental area (29–31), a pathway implicated in curbing drug-seeking (31). Thus, NAC-shell maturation could explain the genetic (10, 11, 13) and epidemiological(61) overlap between impulse-control deficits and obesity, as observed in children with ADHD(16, 17, 19, 20). Because 22q11DS heightens vulnerability to both impulsivity (54–56) and obesity(40, 62), it provides a unique model for investigating shared mechanisms (41). Our results confirmed that deficient NAC-shell maturation predicted higher impulsivity in both HCs and 22q11DS, with dose-dependent effects. Increased frequency and severity of NAC-shell deficits in 22q11DS largely explained impulsivity differences observed across HC and 22q11DS populations, consistent with a shared endophenotype underlying impulsivity and obesity genetic risk. Mediation analyses further showed that NAC-shell maturation fully explained the association between impulsivity and BMI growth across both 22q11DS and HC populations.

Developmentally, BMI deviations linked to atypical NAC-shell maturation were most pronounced during transitions from childhood to adolescence and into early adulthood. This aligns with evidence that the timing of BMI acceleration predicts long-term obesity risk: while adolescent BMI increases are physiological, an early adiposity rebound in childhood signals persistent BMI elevation (4, 35, 36). Mechanisms driving this rebound remain unclear, with most research focusing on environmental factors (4, 35, 36), although neurodevelopmental risk factors (63, 64), such as low birth weight have also been implicated (65). Our findings suggest that impaired NAC-shell inhibitory control may represent a key endogenous pathway linking early BMI acceleration to later obesity. The childhood period also showed the largest impulsivity differences across both HC and 22q11DS NAC-shell subgroups, consistent with ADHD evidence showing that childhood impulsivity precedes and predicts later BMI increases(17, 66). Despite these strong associations, impulsivity followed an inverse U-shaped trajectory, with transient increases during adolescence that could not be fully explained by NAC-shell maturation alone. This pattern fits with the dual-system model(58), which posits that adolescent impulsivity reflects an imbalance between a slowly maturing inhibitory control system (51) and a transient rise in dopaminergic incentive motivation (59). In line with this model, our results suggest that mid-adolescent impulsivity peaks correspond to transient dopaminergic activity (59), while the persistence of impulsive behavior before and after adolescence results from insufficient maturation of top-down inhibitory circuits such as the NAC-shell.

By attenuating NAC-shell maturation, 22q11DS amplifies genetic vulnerability to both impulsivity and maladaptive metabolic trajectories. Yet similar developmental effects appeared in HCs, indicating that this circuit’s role extends to the general population and may explain the link between childhood impulsivity and adult obesity. Neurodevelopmental variance observed within both groups further suggests contributions from polygenic influences beyond the 22q11.2 locus (13). Familial analyses confirmed that NAC-shell maturation variance was shared among siblings and overlapped with familial patterns in impulsivity and BMI, consistent with polygenic risk for inhibitory-control deficits(13). Moreover, NAC-shell maturation directly explained familial co-occurrence of high impulsivity and atypical BMI trajectories, supporting its endophenotypic role in mediating both genetic risk associated with 22q11DS, and additional obesity/impulsivity polygenic risk shared among affected and unaffected siblings. Familial BMI added a smaller, independent effect, possibly reflecting environmental exposure (67) or genetic influences not captured by NAC-shell function. NAC-shell maturation however substantially shaped BMI trajectories regardless of familial BMI, suggesting it may play a central role in amplifying or buffering the effects of both genetic and environmental risk.

This study has several limitations. First, environmental and lifestyle measures were limited, yet such factors likely interact with NAC-shell variability to shape BMI (10). Second, impulsive eating behaviors were not directly assessed, although they were previously linked to obesity in ADHD (26–28) and to NAC-shell dysfunction in animals (22). Moreover, additional downstream consequences of NAC-shell dysfunction may also contribute to this association. For example, NAC-shell’s strong negative connectivity with amygdala and insula recapitulates inhibitory projections observed in animal models and involved in emotional regulation (49). This raises the possibility that NAC-shell dysfunction may promote emotional dysregulation, a core feature of impulsive behavior in ADHD (68–70) and a recognized driver of impulsive eating (71). Finally, while NAC-shell dysfunction is interpreted as a genetically driven endophenotype, additional mediators, such as sleep disturbance, may contribute. Sleep dysfunction is common in 22q11DS (72) and directly linked to impulse-control deficits (73).

Despite these limitations, our findings carry important clinical implications. Assessing NAC-shell maturation could enable early identification of children at risk for chronic obesity, years before BMI deviations emerge. Importantly, impulsivity, measured using a gold-standard CPT neurocognitive battery, did not fully capture the obesity risk associated with atypical NAC-shell maturation, suggesting that NAC-shell metrics may provide additional clinical value for identifying vulnerable youth. While fMRI is not feasible for large-scale screening, our findings could inform the development of assessment batteries that more directly probe NAC-shell function(74, 75), including through smartphone-based behavioral and neuropsychological paradigms designed for scalable deployment(76, 77). These tools could be implemented in routine pediatric settings to support early obesity-risk stratification and targeted prevention efforts.

From a preventive perspective, animal studies clearly indicate that weight gains associated with NAC-shell circuit dysfunction are tightly modulated by the quantity and quality of immediately available food options(22). As such, interventions that modify environmental food exposure may be especially effective for individuals with immature NAC-shell function, who may struggle to inhibit impulsive eating despite acute awareness of its consequences. This underscores the need for societal strategies targeting food availability(78, 79), for instance, through school nutrition programs(80, 81), which might prove more effective than relying solely on awareness campaigns(82). Beyond environmental interventions, approaches enhancing impulse control such as targeting dysregulated sleep patterns (83) linked to both impulsivity (84–86) and obesity (87), may strengthen NAC-shell function and mitigate obesity risk(83).

Finally, our findings may clarify mechanisms underlying current pharmacological treatments. Psychostimulants used for ADHD, for example, improve both impulsivity(88) and obesity risk (16, 89), with some evidence of differential effects of amphetamines versus methylphenidate on weight(90). Investigating whether these differences reflect distinct influences on NAC-shell function could inform personalized treatments. Similarly, it will be important to assess whether GLP-1–based obesity treatments also impact impulsivity by modulating NAC-shell circuitry behavior (91). More broadly, it would be impart to investigate whether NAC-shell dysfunction may represent a transdiagnostic endophenotype for impulse-control deficits observed across a range of psychiatric conditions (92), including borderline personality disorder (93), conduct disorder (94), substance use disorder (95) and bipolar disorder (96), all of which share overlapping genetic risk architectures (13).

In summary, we identified NAC-shell inhibitory circuit maturation as a shared neurodevelopmental mechanism linking childhood impulsivity to obesity risk. By bridging genetic vulnerability, behavioral regulation, and metabolic outcomes, NAC-shell maturation emerges as a key endophenotype for understanding and potentially mitigating obesity and related impulsivity-driven conditions.

## 4. Methods

### 4.1 Participants

Two independent samples of Healthy Controls (HCs) were recruited for the purpose of NAC-shell segmentation and validation. A first HC discovery sample (HC-ds) was recruited through the Generva-22q11DS Longitudinal Study mostly among unaffected siblings of 22q11DS participants (107/136) (97, 98), and was composed of a total of 136 participants (M/F=62:74) followed up for a total of 227 longitudinal assessments. From this HC-ds NAC-shell segmentation analysis was performed in a cross-sectional subsample of 55 adult participants (M/F=28:28, Mean-Age=22.73+/− 3.47). NAC-shell segmentation results were then validated in a second independently recruited sample of 55 healthy controls adult (M/F=30:25) recruited through web announcements and local database in the context of a study carried out at Mood clinic of the Psychiatry Department of Geneva University Hospital (99). Subsequently normative trajectories of NAC-shell maturation were estimated in the entire longitudinal HC-DS sample as described below. Finally, a sample of 126 individuals with 22q11DS (M/F=67:59) were recruited through the Geneva-22q11DS Longitudinal Study(98, 100) and followed up for a total of 233 longitudinal assessments. All participants signed a written informed consent (ethical approval from Geneva University CER 13-081).

### 4.2 Functional MRI Acquisition, Processing

In both HC-ds and 22q11DS samples Resting-State functional MRI (RS-fMRI) scans were obtained using a T2-weighted sequence with the following parameters: 200 frames, voxel size=1.84×1.84×3.2mm^3^, repetition time=2400ms. Scanning sessions lasted 8 minutes and were preprocessed using an in-house pipeline described in previous publications in both HC-ds and 22q11DS samples, Resting-State functional MRI (RS-fMRI) scans were obtained using a T2-weighted sequence with the following parameters: 200 frames, voxel size□=□1.84×1.84×3.2□mm³, repetition time□=□2400□ms. Scanning sessions lasted 8 minutes during which, participants were instructed to fixate on a white cross presented on a black screen and to remain awake. Structural images were acquired using a T1-weighted sequence with the following parameters: 0.86□×□0.86□×□1.1□mm³ resolution, 192 coronal slices, repetition time□=□2500□ms, echo time□=□3□ms, acquisition matrix□=□224□×□256, field of view□=□22□cm², and flip angle□=□8°. Images were preprocessed using an in-house pipeline described in previous publications. (37, 101). In the HC-rs samples RS-fMRI scans were obtained with 3T Siemens scanner equipped with a 32-channel head coil at the Brain & Behavior Laboratory, University of Geneva. Rs-fMRI was obtained by a T2*-weighted echo-planar imaging (EPI) sequence. A total of 250 functional volumes were collected, each consisting of 36 axial slices (TR□=□2100□ms, TE□=□30□ms, flip angle□=□80°, field of view□=□205□mm, matrix size□=□64□×□64, voxel size□=□3.2□mm³ isotropic, distance factor□=□20%). High-resolution anatomical images were also acquired using a T1-weighted sequence (TR□=□1900□ms, TI□=□900□ms, TE□=□2.27□ms, flip angle□=□9°, field of view□=□256□mm, matrix size□=□256□×□256, slice thickness□=□1□mm³, 192 sagittal slices, phase encoding direction: anterior-to-posterior). Four dummy scans (∼9□s) were included at the beginning of each fMRI run. During scanning, participants were instructed to stay awake, breathe normally, and let their mind wander without focusing on anything for a total duration of 8 minutes.

### 4.3 Statistical Analyses

#### 4.3.1 Functional Dissection of NAC circuits in HC-Discovery and Replication Samples

We then employed a recently published data-driven μCAPs algorithm to dissect functionally distinct Nucleus Accumbens (NAC) circuits, characterized by distinct anatomical gradients with the NAC seed, mapping onto distinct whole brain co-activation patterns. Briefly, this method extends conventional co-activation pattern (CAP) analysis by accounting for spatial heterogeneity within the seed region(37). Through an iterative clustering process based on the dynamic BOLD signal, μCAPs identifies temporally recurring and spatially distinct subpatterns of seed co-activation with the rest of the brain. This approach enables a fine-grained functional dissection of NAC circuits based on heterogenous dynamic connectivity profiles of its different components, providing insights into NAC functional subdivisions beyond anatomical priors. NAC functional circuit dissection was performed in a cross-sectional subsample of 56 adult participants (M/F=28:28, Mean-Age=22.73+/− 3.47) recruited in the HC discovery sample.

We validated the consistency of μCAPs functional dissection of NAC circuits in an independent adult HC replication sample. Within this sample, BOLD activity from the entire NAC seed was averaged, z-scored, and thresholded at 0.5 to identify time points with maximal NAC activity, in accordance with the initialization step of the μCAPs algorithm. Frames with significant NAC activity were then assigned to the closest matching NAC coactivation pattern defined in the HC discovery sample. Importantly, such matching was performed based exclusively on voxel-wise activity outside the NAC seed region. We then tested whether frames matched exclusively according to extra-seed coactivation patterns yield similar activation gradients within the NAC seed, providing evidence of consistency in NAC segmentation results across independent samples.

#### 4.3.2 Definition of NAC-shell Circuit Maturation Patterns in Longitudinal HC and 22q11DS Samples

Having validated the μCAPs functional dissection of the candidate NAC-shell inhibitory control-circuit, we then investigated the circuits normative developmental trajectory in the full longitudinal HC sample. To this end, we once again identified frames with significant NAC activity, which we matched with co-activation patterns defined in the adult HC discovery sample. For each longitudinal visit, we averaged frames assigned to each adult NAC segment. We then investigated how such co-activation patterns evolved with age, focusing specifically on the segment that most closely resembled previous anatomical segmentation of the Shell portion of the NAC, here defined as NAC-shell. We used a previously published multivariate Partial-Least-Square (PLS) approach (https://github.com/valkebets/myPLS-1)(101, 102) to analyze how NAC-shell co-activation patterns changed as a function of age in the longitudinal HC sample. The brain-loadings resulting from this normative PLS analysis were then used to generate a NAC-shell Maturation Score (NAC-shell-MS) through matrix with the NAC-shell co-activation pattern observed at individual longitudinal assessments. We then modelled such NAC-shell-MS as function of age through Mixed-Model-Linear-Regression (MMLR) (103, 104), comparing male and female HC individuals. Brain loadings derived from the normative PLS analysis performed on the HC sample, were also employed to derive NAC-shell-MS in the 22q11DS sample. We compared HC and 22q11DS NAC-shell-MS with MMLR, to characterize differences in NAC-shell maturation across groups.

We employed normative age-related NAC-shell PLS analysis to characterize behavioral correlates of typical and atypical NAC-shell maturation. Specifically, we computed the difference between NAC-shell-MS measured at each longitudinal assessment in both HC and 22q11DS and the age-matched and gender-specific normative NAC-shell-MS trajectories defined in the HC sample. We averaged such measure across all longitudinal assessments to estimate whether NAC-shell-MS of an individual was on average higher or lower than the normative trajectory, which we used to define High/Typical-vs-Low/Atypical NAC-shell-MS subgroups in both HC and 22q11DS populations. See Table 1 for demographics of subgroups. Differences in the proportion of individuals classified as having atypically low NAC-shell-MS across samples were used to estimate the impact of 22q11DS-related genetic vulnerability on circuit maturation. Moreover, by comparing typical versus atypical NAC-shell subgroups within both cohorts, we evaluated transdiagnostic effects on obesity- and impulsivity-related phenotypes.

**Table 1:**
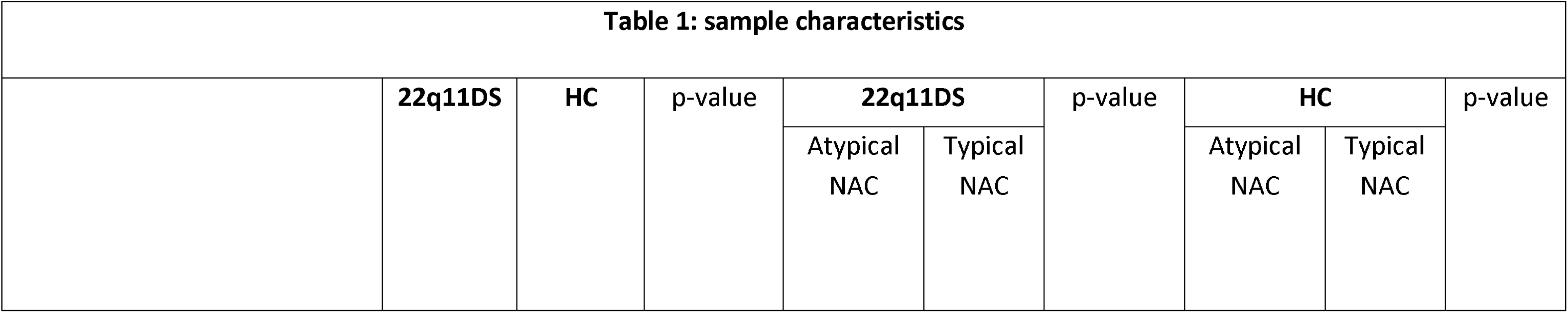
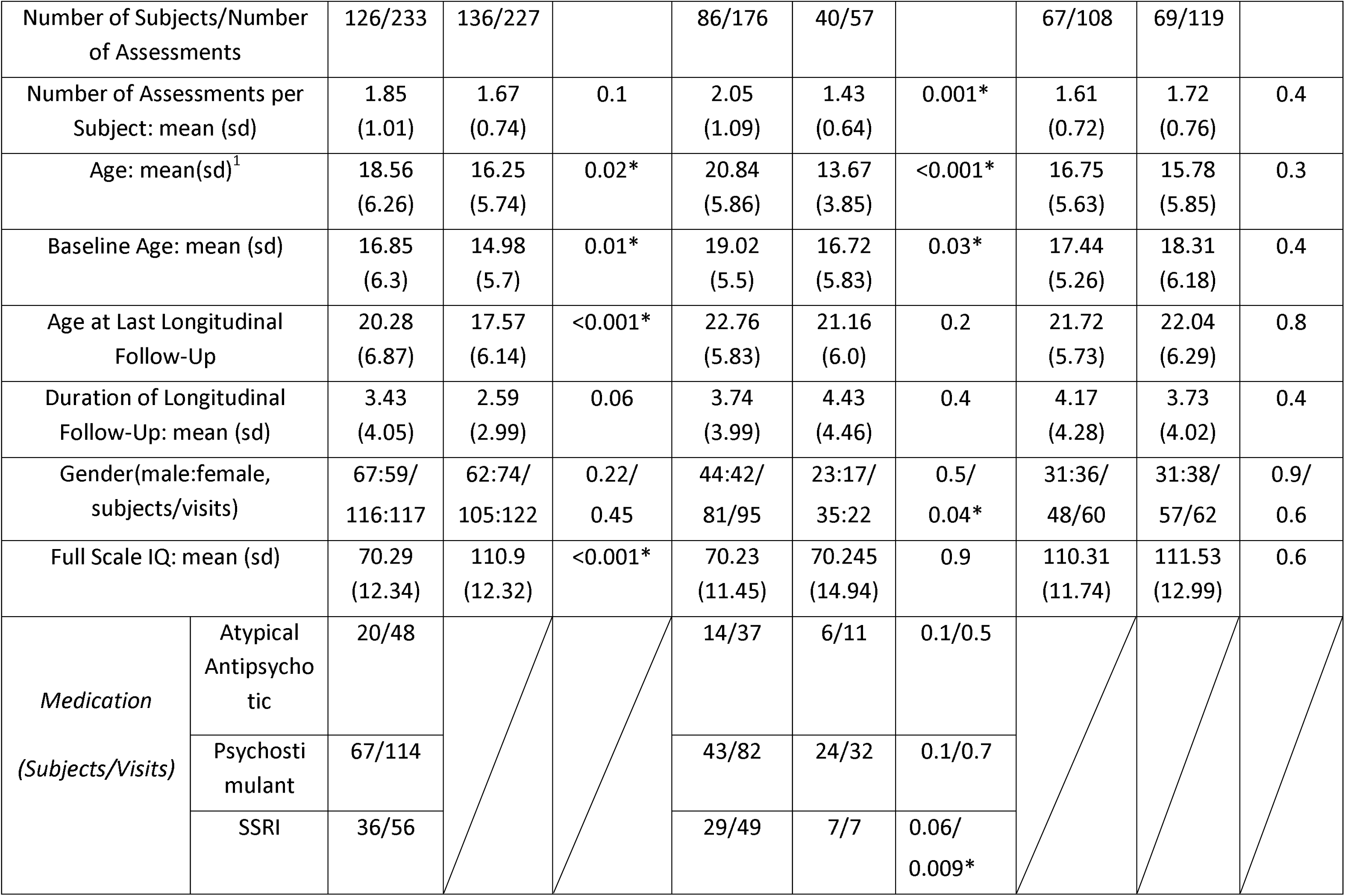
Demographic information. Differences in continuous measures were tested using a two-samples t-test, while binary measures were tested using a Chi-squared test. ^1^Two-samples t-test ^2^Chi-squared test *Significant difference (p<0.05)

#### 4..3.3 Behavioral Correlates of NAC-shell Maturation in HC and 22q11DS samples

We compared BMI trajectories across NAC-shell subgroups using MMLR, both within 22q11DS and HC diagnostic categories, and through a 4-group transdiagnostic analysis, testing if a transdiagnostic gradient of NAC-shell Maturation, going from High/Typical-HC to Low/Atypical-22q11DS subgroups, would be reflected in a similar transdiagnostic gradient of BMI trajectories. We included only participants who were naïve to antipsychotic medication. We further investigated whether NAC-shell-MS was directly correlated with differential longitudinal BMI trajectories. To this end we compared each subjects’ BMI longitudinal rate of change with the age-matched normative BMI trajectory deriving a Longitudinal-BMI-Trajectory score for each subject. We computed Spearman correlations between Longitudinal-BMI-Trajectory score and NAC-shell-MS score, across the entire sample and within 22q11DS and HC populations.

We then explored a possible relationship between NAC-shell-MS and neurocognitive developmental trajectories of attention and inhibitory control as estimated by the Conners’ Continuous Performance Test 2^nd^ CPT2 or 3^rd^ edition, CPT3(105, 106). Use of different CPT versions did not differ across groups. See Table 1. We corrected the effects of version with linear regression. Previous literature demonstrated that CPT scores are differentially influenced by different neurocognitive dimensions, which can be captured through multivariate latent structure analysis (53). We hence employed multivariate PLS analysis to characterize normative developmental trajectories of such underlying neurocognitive dimensions using the same analysis pipeline employed for NAC-shell-MS. We first performed a normative PLS analysis to characterize the effects of age and gender on CPT measures of omission and commission errors, detectability, Hit-Reaction-Time (HRT), perseveration errors, HRT standard deviation, and variability in the HC sample. Loading of CPT variables derived from such normative PLS analysis were used to derive normative CPT-Maturation-Scores (CPT-MS) at each longitudinal assessment in both the HC and 22q11DS samples. We then compared such CPT-MS across male and female HC individuals, across HC and 22q11DS populations, and across NAC-shell subgroups using MMLR. Additionally, we computed Spearman correlations between average NAC-shell-MS and average CPT-MS.

Finally, we explored the possible 3-way interaction between NAC-shell, CPT and BMI trajectories. We specifically tested whether NAC-shell-MS mediated the relationship between CPT-MS and longitudinal BMI-Trajectory-Score. Mediation analyses were conducted in MATLAB employing a linear model framework with bootstrap inference. (details in Supplementary Materials).

#### 4.3.4 Genetically informed mediation analyses

We conducted a series of analysis to explore genetic factors influencing NAC-shell, CPT and BMI trajectories. We firstly tested whether NAC-shell-MS mediated the effects of 22q11DS diagnosis on CPT-MS and longitudinal BMI-Trajectory-Scores.

We further explored the influence of familial genetic factors beyond the 22q11.2 locus, by measuring familial phenotyping correlations among siblings. Specifically, we calculated Spearman correlations between an individual’s NAC-shell-MS, and the mean corresponding scores of his siblings across 53 families (mean family size 2.4 ± 0.63 siblings), including 54 individuals with 22q11DS and 73 unaffected siblings. Parallel correlations were computed for BMI-Trajectory Scores (32 families, mean = 2.5 ± 0.72) and CPT-Impulsivity-MS (51 families, mean = 2.4 ± 0.63). Was also computed across trait correlations between familial and subject-level NAC-shell-MS, BMI-Trajectory Scores and CPT-Impulsivity-MS.

We then conducted a total of 5 mediation analyses to explore pathways linking familial and subject level NAC-shell-MS and BMI-Trajectory scores to each other. We firstly conducted 3 mediation analyses to test whether the effects of Familial-CPT-Impulsivity-MS, Familial-NAC-shell-MS and Familial-BMI-Trajectory-Scores on Self-BMI-Trajectory-Score were mediated by Self-NAC-shell-MS. Given significant associations observed between Familial-BMI-Trajectory-Scores and Familial-NAC-shell-MS, we additionally tested if the association between Self-NAC-shell-MS and Familial-BMI-Trajectory-Scores was mediated by Familial-NAC-shell-MS and whether Familial-BMI-Trajectory-Scores contributed to mediate the association between Familial-NAC-shell-MS and Self-BMI-Trajectory-Score.

Finally, to evaluate the combined impact of neurodevelopmental and familial factors on obesity risk, we grouped participants according to both NAC-shell maturation status (typical vs. atypical) and familial BMI trajectory (low vs. high). We tested the cumulative effects of Family-BMI and Self-NAC by modelling longitudinal BMI trajectory across the 4 resulting subgroups (Typical-NAC/Low-Family-BMI, Typical-NAC/High-Family-BMI, Atypical-NAC/Low-Family-BMI, Atypical-NAC/High-Family-BMI) using linear mixed-effects models (BMI ∼ Group + Age * Group + (1|ID)). We also conducted pairwise comparisons of Self-BMI-Trajectory scores across the 4 subgroups using pairwise Wilcoxon rank-sum tests.

## 5. Acknowledgments

This study was supported by the Swiss National Science Foundation (SNSF) (Grant numbers: to SE FNS 320030_179404, FNS 324730_144260) and by the National Center of Competence in Research (NCCR) Synapsy-The Synaptic Bases of Mental Diseases (SNF, Grant number: 51AU40_125759). MarS (#163859), MauS (#162006) and CorrS (#209096) were supported by grants from the SNF. Dr. Corrado Sandini also received support from the Fondation Gertrude von Meissner, grant no. 52, March 2025. Samuele Cortese, NIHR Research Professor (NIHR303122) is funded by the NIHR for this research project. The views expressed in this publication are those of the author(s) and not necessarily those of the NIHR, NHS or the UK Department of Health and Social Care. Samuele Cortese is also supported by NIHR grants NIHR203684, NIHR203035, NIHR130077, NIHR128472, RP-PG-0618-20003 and by grant 101095568-HORIZONHLTH- 2022-DISEASE-07-03 from the European Research Executive Agency.

We warmly thank all the families who participated in the study.

## 6. Disclosure

Prof. Cortese has declared reimbursement for travel and accommodation expenses from the Association for Child and Adolescent Central Health (ACAMH) in relation to lectures delivered for ACAMH, the Canadian AADHD Alliance Resource, the British Association of Psychopharmacology, Healthcare Convention and CCM Group team for educational activity on ADHD, and has received honoraria from Medice. All the other authors declare that they have no conflict of interest. Valeria Parlatini is recipient of an NIHR Advanced Fellowship (NIHR305518). *The views expressed are those of the author(s) and not necessarily those of the NIHR or the Department of Health and Social Care. VP also received travel and fees reimbursement by Medice to attend Eunethydis 2025,* none of this related to the present project.

## 7. Data Availability

The datasets generated during and/or analysed during the current study are not publicly available due to privacy or ethical restrictions on human participant data. De-identified data may be made available to qualified researchers upon reasonable request to the corresponding author, subject to approval by the relevant ethics board.

## 8. Author Contribution Statement

*C.S., F.D. and S.E* conceived and designed the study.

*F.D. D.VDV, C.S and N.R.* developed the methodology.

*A.I, C.L, S.E, M.SH, L.S, C.P and S.E* performed the experiments and collected the data.

*C.S, F.D, N.R, S.F and N.K* conducted the data analysis and interpreted the results

*C.S* drafted the manuscript with input from *N.R, D.VDV, S.E, S.C, and V.P*.

*S.E. S.E M.SH, C.P and D.VDV*. supervised the project.

All authors critically revised the manuscript for important intellectual content and approved the final version for publication.

All authors agree to be accountable for all aspects of the work.

## Notes

### Author Declarations

Ethics committee Commission Cantonale d'Ethique de la Recherche sur l'etre humain of Canton de Geneve gave ethical approval for this work

