## Supplementary Material for "Blunted maturation of inhibitory control circuits in the NAC-shell underlies genetic vulnerability to early-life obesity and impulsivity"

1 Table of Contents

### 2  Functional MRI Acquisition, Processing

In both the Healthy Control Discovery Sample (HC-DS) and 22q11.2 Deltion Syndrome (22q11DS) samples Resting-State functional MRI (RS-fMRI) scans were obtained using a T2-weighted sequence (200 frames) with following parameters: acquisition matrix = 94×128, field of view = 96x128, voxel size = 1.84×1.84×3.2 mm3, 38 axial slices, slice thickness = 3.2 mm, TR=2400 ms, TE=30 ms, flip angle=85°, phase encoding A >> P, descending sequential ordering, GRAPPA acceleration mode with factor PE=2. RS-fMRI data were processed using SPM12 (Wellcome Trust Centre for Neuroimaging, London, UK: <http://www.fil.ion.ucl.ac.uk/spm/>) and the Data Processing Assistant for rs-fMRI (DPARSF) [1]. For each participant, functional images were realigned over time and spatially smoothed with an isotropic Gaussian kernel of 3 mm full width at half maximum (FWHM). Subsequently, anatomical images were coregistered to the functional space and segmented with the SPM12 Segmentation algorithm [2], Nuisance variables were regressed out (6 head motion parameters + other 6, average cerebrospinal fluid, and white matter signal). The first five frames were excluded from the analysis to account for magnetization equilibration. The time series were linearly detrended and translational and rotational movement parameters as well as global, mean white matter and mean cerebrospinal fluid signals were regressed out from the BOLD time series using the warped DPARSF[1] tissue masks. After preprocessing, all functional images were warped to the DARTEL space, and then to MNI space to create population-based results. Extended motion correction through scrubbing of frames with high framewise displacement (FD >0.3 mm; [3]) is included within the μCAPs algorithm to exclude them from the analysis.

### 3 μCaps functional dissection of NAC circuits in NT-Discovery Sample

#### 3.1 Methods

We then employed a recently published data-driven μCAPs algorithm [4] to identify distinct Nucleus Accumbens (NAC) circuits characterized by distinct anatomical gradients within the NAC seed region, mapping onto differential whole-brain functional co-activation patterns. (code for this process is available at: <https://github.com/MIPLabCH/mCAP>)

The aim of the method was to identify K μCAPs where each μCAP is also associated with a weight map for the seed ROI, in this case the Nucleus Accumbens. Starting with an initial seed weight map, for each, the corresponding time course is computed. These time courses are subsequently Z-scored over time, as per standard co-activation pattern (CAP) analysis procedures. This allows each time course to reflect the temporal activity patterns associated with the seed maps. For each seed map, fMRI volumes are tagged as “selected” if the Z-scored time course exceeds a certain threshold. In the initial iteration, a threshold of 0.5 is applied to enhance sensitivity. For subsequent iterations, the threshold is adjusted to 1.0, a common choice in conventional CAP analysis[5, 6]. The selected fMRI frames across all seed maps are then clustered using K-means clustering with a 1-correlation distance metric. The optimal number of clusters, K, is determined, and a representative centroid for each cluster is calculated by averaging the frames assigned to it. These centroids are referred to as micro-co-activation patterns (μCAPs). To update the seed weight maps, each μCAP is restricted to the seed region, applying a "winner-takes-all" approach. Specifically, for each voxel, only the μCAP with the highest value is retained, while values from other μCAPs are set to zero. This updated set of seed weight maps is then utilized in the next iteration of the algorithm. Convergence is monitored by calculating the distance between μCAP seed maps across successive iterations. Seed maps from consecutive iterations are matched using the Hungarian algorithm[7], and the convergence threshold is defined as an average cosine distance of 0.005. The initial iteration, based on a uniform seed pattern, is excluded from the convergence assessment.

#### 3.2  Results

The μCAPs approach identified **six distinct** NAC circuits, each characterized by distinct anatomical activation gradients with the NAC seed region mapping onto unique whole-brain functional coactivation patterns. Supplementary Figure 1 provides a visual representation of both within-seed and whole-brain coactivation patterns for each CAP, while the following section provides a brief anatomical description of these patterns.

**CAP-1** was identified as a candidate **NAC-shell circuit**.
Its within-seed coactivation pattern was centered in the **ventral–posterior–inferior** portion of the NAC and was anatomically consistent with previous histological [1] and structural connectivity-based [2,3] segmentations of the NAC shell. The whole-brain coactivation pattern showed **positive coactivation** with higher-order frontoparietal cortices and the **anterior hypothalamus**, while **negative coactivations** were observed in the **posterior–lateral hypothalamus**, **VTA**, **insula**, **amygdala**, and **medial temporal cortices**.

**CAP-2** exhibited within-seed coactivation centered in the **dorsal–posterior** portion of the NAC.
Its whole-brain coactivation pattern was characterized by **widespread positive coactivation** across the **inferior–lateral and medial frontal cortices**, **cuneus**, **precuneus**, and **insula**, along with **subcortical coactivation** in the **dorsal striatum**, **thalamus**, and both **anterior and posterior–lateral hypothalamus**. **Negative coactivations** were observed in the **VTA**, **orbitofrontal cortex**, and **inferior temporal cortex**.

**CAP-3** showed within-seed coactivation centered in the **ventral–posterior** portion of the NAC.
Its whole-brain coactivation pattern included **positive coactivation** in the **ventromedial and superior frontal regions**, **precuneus**, and **inferior and medial temporal cortices**, as well as in **subcortical regions** including the **anterior hippocampus**, **anterior caudate**, and **VTA**. **Negative coactivations** were observed in the **midcingulate cortex**, **thalamus**, **opercular**, and **central occipital regions**.

**CAP-4** displayed within-seed coactivation centered in the **ventral–posterior** NAC, resembling that of CAP-3. However, its whole-brain coactivation pattern was distinguished by **positive coactivation** in the **dorsal midcingulate**, **primary motor cortex**, **insula**, **thalamus**, **visual cortex**, and **medial parietal regions**, along with **subcortical coactivation** in the **VTA** and **hypothalamus**. **Negative coactivations** were found in the **medial prefrontal** and **lateral parietal cortices**, as well as the **posterior cingulate/precuneus**.

**CAP-5** showed within-seed coactivation strongest in the **central portion** of the NAC.
It displayed **positive coactivation** with the **medial prefrontal cortex**, **posterior cingulate/precuneus**, and **lateral parietal cortex**, as well as **subcortical coactivation** in the **VTA** and **hypothalamus**. **Negative coactivation** was observed in the **frontoparietal executive cortices**.

**CAP-6** showed within-seed coactivation strongest in the **central and ventral portions** of the NAC.
It was characterized by **widespread positive coactivation** across most cortical and subcortical regions, with peaks in the **midcingulate cortex** and **paracentral gyrus**, while **negative coactivations** were observed in the **visual** and **ventral frontal cortices**.

Supplementary Figure 1: μCaps functional dissection of NAC circuits in Adult Healthy Controls

Each line represents one of six circuits derived from data driven Caps segmentation of the Nucleus Accumbens (NAC). Brain plots represent the average brain activation patterns of frames assigned to each NAC segment. Columns on the right depict the activation pattern within the NAC seed, ,describe which anatomical portion of the NAC contributed to the overall seed activation pattern within selected frames, with red-yellow shading indicating stronger activations. For Whole-Brain Co-Activation-Patterns (CAPs), Red-yellow shading indicates positive co-activations with the corresponding with portion of the NAC. Blue shading indicates negative co-activations, signifying that the corresponding brain region had lower than average activations in frames when the corresponding anatomical portion of the NAC had highest activity. CAP1 had a within seed pattern that was anatomically consistent with previous attempts at NAC-shell segmentation, with highest activity in the ventral and anterior portions of the NAC. Moreover, CAP1 brain pattern was uniquely characterized by strong positive co-activations in higher-order fronto-parietal cortices, including in particular Dorso-Lateral-Prefrontal-Cortex (highlighted in pink) combined with negative co-activations with both Ventral-Tegmental-Area (VTA Highlighted in Green), and Posterior-Lateral portions of the Hypothalamus (Highlight in Yellow). CAP1 pattern is therefore highly consistent with functional role of NAC-shell in mediating top-down Prefrontal inhibitory controls of both VTA-mediated goal-directed behavior and of feeding behavior mediated by Posterior-Lateral-Hypothalamus.

### 4 Validation of μCaps functional dissection of NAC circuits across HC-Discovery and HC-Replication Samples

#### 4.1 Methods:

Results of μCAPS functional dissection of NAC circuits were validated in a second, independently recruited sample of 55 healthy adults (M/F = 30/25) recruited through web announcements and a local database, within the framework of a study conducted at the Mood Clinic of the Psychiatry Department, Geneva University Hospital [8].

For this sample, BOLD activity from the entire NAC seed was averaged, z-scored, and thresholded at 0.5 to identify time points of maximal NAC activation, consistent with the initialization step of the μCAPs algorithm. Frames with significant NAC activity were then assigned to the closest matching NAC coactivation pattern identified in the HC discovery sample. Importantly, this matching was performed based exclusively on voxel-wise activity **outside** the NAC seed region (Supplementary-Figure-1-Panel-1). We then assessed whether frames matched solely according to extra-seed coactivation patterns yielded similar activation gradients within the NAC seed, thus providing evidence for the consistency of NAC segmentation across independent samples (Supplementary-Figure-1-Panel-2). Specifically, we estimated the correlation between NAC-seed coactivation patterns across the discovery and replication samples for frames matched exclusively based on extra-seed coactivation patterns. Furthermore, we tested whether the average correlation between NAC-seed coactivation patterns for matched CAPs was significantly higher than a random distribution derived from 1000 random matchings between discovery and replication samples (Supplementary-Figure-1-Panel-2C).

#### 4.2 Results:

NAC circuits identified by the μCAPs approach were highly consistent across adult HC-Discovery and HC-Replications samples, both in terms of within-seed and whole-brain coactivation patterns, across each of the 6 NAC circuits (See Supplementary Figure 2). Indeed, Correlations between extra-seed whole-brain coactivation patterns used to match CAPs between the discovery and replication samples averaged **r = 0.85**, ranging from **0.75 to 0.95** (Supplementary-Figure-1-Panel-1).

Correlations between within-seed NAC coactivation patterns were highly significant across all six CAPs, ranging from **r = 0.58 to 0.85** (Supplementary-Figure-1-Panel-2A-2B). The average correlation for matched CAPs was **r = 0.73**, which was significantly higher than the null distribution obtained from 1000 random matchings **(mean = 0.65 ± 0.03, p = 0.0006**; Supplementary-Figure-1-Panel-2C).

Given that CAPs were matched independently of NAC seed activity, the observed similarity strongly supports the replicability of the μCAPs-based NAC circuit segmentation across the discovery and replication samples.

Supplementary Figure 2: Comparison μCAPs NAC circuits across HC-Discovery and HC-Replication Samples

Each line represents one of six circuits derived from data driven μCaps segmentation of the Nucleus Accumbens (NAC). Columns on the left depict NAC circuits identified in the adult Healthy Control (HC) Discovery Sample while column on the right depict results in the HC-Replication-Sample. Brain plots represent the average brain activation patterns of frames assigned to each NAC segment. Activation pattern within the NAC seed, describe which anatomical portion of the NAC contributed to the overall seed activation pattern within selected frames, with red-yellow shading indicating stronger activations. For Whole-Brain Co-Activation-Patterns (CAPs), Red-yellow shading indicates positive co-activations with the corresponding with portion of the NAC. Blue shading indicate negative co-activations, signifying that the corresponding brain region had lower than average activations in frames when the corresponding anatomical portion of the NAC had highest activity.

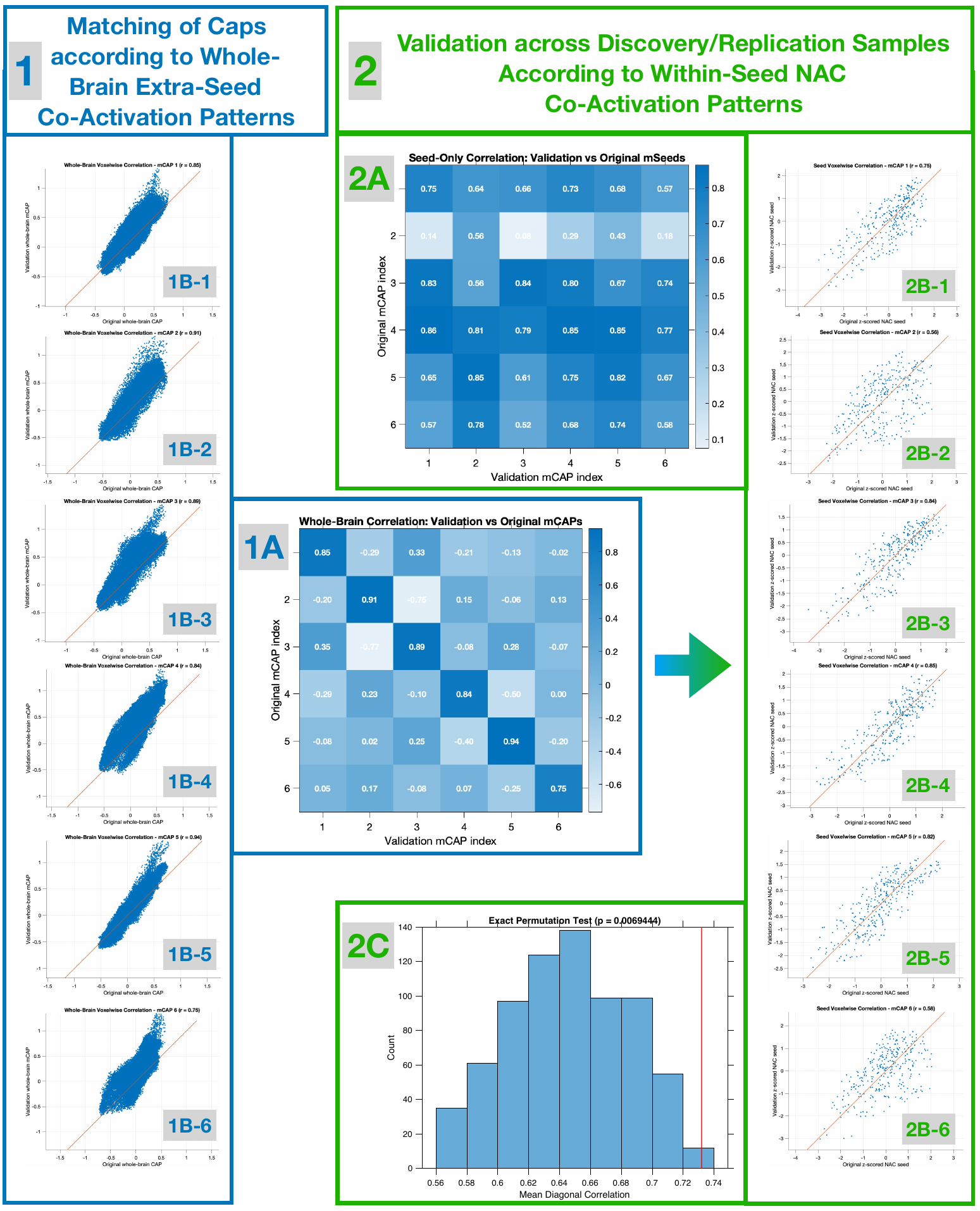

Supplementary Figure 3: Statistical validation of μCAPs NAC functional circuit consistency across HC-Discovery and HC-Replication Samples.

**Panel 1:** Correlation of whole-brain extra-seed coactivation patterns for each of the six CAPs. **Panel 1A:** Adjacency matrix showing correlations between coactivation patterns of corresponding CAPs across the discovery and replication samples (diagonal elements), and correlations between non-matched CAPs (off-diagonal elements). **Panel 1B:** Scatter plots illustrating voxel-wise similarity in whole-brain coactivation patterns across samples for each of the six matched CAPs (Panels 1B-1 to 1B-6). **Panel 2:** Correlation of within-seed NAC coactivation patterns for each of the six CAPs. **Panel 2A:** Adjacency matrix showing correlations between NAC coactivation patterns of corresponding CAPs across the discovery and replication samples (diagonal elements), and correlations between non-matched CAPs (off-diagonal elements). **Panel 2B:** Scatter plots illustrating voxel-wise similarity in within-seed NAC coactivation patterns across samples for each of the six matched CAPs (Panels 2B-1 to 2B-6).

### 5 Neurocognitive Correlates of NAC-shell Maturation – Attentional Component

Here, we report the trajectory of the CPT-Attentional component estimated by the normative PLS (see maintext). This first component (R=0.64, p=0.001, Supplementary-Figure-4-Panel-A), captured **progressive improvements in attention performance reflected by reductions in omission, commission, and perseveration errors, improved detectability, and decreased Hit Reaction Time (HRT) and variability**. The corresponding **CPT-Attention Maturation Score (CPT-Attention-MS)** increased sharply through childhood and early adolescence and plateaued in early adulthood (Supplementary-Figure-4-Panel-A2). **This score was lower in 22q11DS regardless of age** (P-Group-Effect=0.005, P-Age-Interaction=0.4, R=0.64, p=0.001, Supplementary-Figure-4-Panel-A3). The CPT-Attentional trajectory was not impacted by the NAC-shell grouping in the HCs (P-Group-Effect=0.86, P-Age-Interaction=0.71, Supplementary-Figure-2-Panel-B2). In 22q11DS sample, there was a mild blunting in trajectory (P-Group-Effect=0.051, P-Age-Interaction=0.021, Supplementary-Figure-2-Panel-B3). This contributed to a small positive association between the average CPT-Attention-Maturation-Score and the average NAC-shell-MS (rho=0.15, p=0.015, Supplementary-Figure-2-Panel-B4), which was however mainly driven by the 22q11DS sample (rho = 0.16, p=0.09) and not observed in the HCs sample (rho = 0.04, p=0.65).

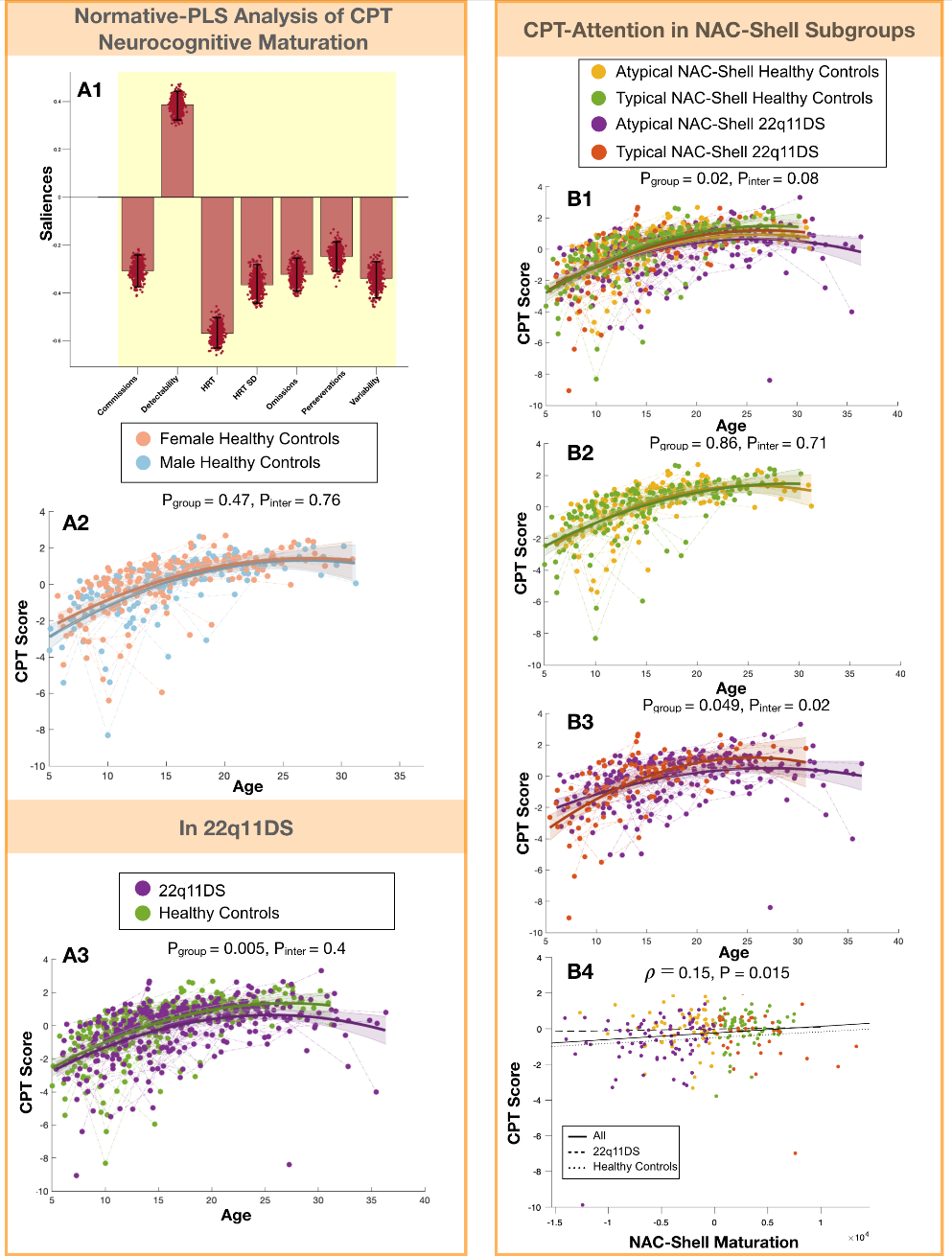

Supplementary Figure 4: Neurocognitive Correlates of NAC-shell Maturation

**Panel A**: Normative-PLS analysis of CPT neurocognitive maturation. **Panel A1**: Multivariate CPT patterns describing contribution of each variable to the corresponding neurocognitive dimension. Variables highlighted in yellow contribute significantly to the pattern. Direction of the bar reflects direction of association (upwards for positive association and downwards for negative association). **Panel A2**: Developmental trajectories CPT-Neurocognitive-Maturation-Score, derived from normative PLS analysis, compared across genders in HCs . **Panel A3**: Developmental trajectories CPT-Neurocognitive-Maturation-Score compared across HCs and individuals with 22q11DS. **Panel B**: CPT-Attention in NAC-shell subgroups. **Panel B1**: Trajectory of CPT-Attention-MS compared across transdiagnostic NAC-shell subgroups**. Panel B2**: Trajectory of CPT-Attention-MS compared across Typical and Atypical NAC-shell Maturation subgroups in HCs**. Panel B3:** Trajectory of CPT-Attention-MS compared across Typical and Atypical NAC-shell Maturation subgroups in 22q11DS. **Panel B4**: Association with Average NAC-shell-Maturation-Score with average CPT-Attention-Maturation-Score.

### 6 Genetically informed mediation analyses

#### 6.1 Methods:

Mediation analyses were conducted using a custom MATLAB script implementing linear models via the *fitlm* function. This framework enabled estimation of both direct and indirect effects. Each mediation analysis consisted of three regression models: (1) the mediator was regressed on the independent variable to assess path *a* (Mediator ~ IndependentVariable); (2) the dependent variable was regressed on both the mediator and the independent variable to estimate paths *b* and *c′* (DependentVariable ~ Mediator + IndependentVariable); and (3) the total effect (path *c*) was evaluated by regressing the dependent variable on the independent variable alone (DependentVariable ~ IndependentVariable). The indirect effect was quantified as the product of paths *a* and *b* (a × b). Statistical significance of the mediation was assessed via nonparametric bootstrapping (1000 iterations), and 95% confidence intervals were derived from the bootstrap distribution. Indirect effects were deemed significant when the confidence interval did not include zero. Results of the mediation analyses are detailed in **Supplementary Table 2**. For each model, Path a represents the effect of the input variable on the mediator, Path b represents the effect of the mediator on the outcome while controlling for the input, Path c represents the total effect of the input on the outcome, and Path c′ represents the direct effect of the input on the outcome after accounting for the mediator. The product of a × b estimates the statistical significance of the mediating effect of the mediator on the association between the input and the outcome.

Having observed robust transdiagnostic associations between NAC-shell maturation and both impulsivity and deviant BMI trajectories, a first mediation analysis **(Model-1)** tested whether atypical NAC-shell maturation mediated the link between these two phenotypes. Next, we conducted genetically informed mediation analyses to explore the role of NAC-shell maturation as a candidate endophenotype mediating the effects of genetic vulnerability on both impulsivity and early-life obesity. We first focused on the genetic vulnerability linked to 22q11DS. Specifically, we conducted two separate mediation analyses testing whether the effects of 22q11DS diagnosis on increased CPT-Impulsivity-MS **(Model-2)** and increased BMI-Trajectory Scores **(Model-3)** were mediated by reductions in NAC-Shell-MS.

Next, we examined mechanisms contributing to significant variance in NAC-Shell-MS observed within both 22q11DS and HC samples, which produced remarkably similar transdiagnostic effects on CPT-Impulsivity-MS and BMI-Trajectory Scores in both groups. We explored the influence of familial genetic factors beyond the 22q11.2 locus by measuring familial phenotypic correlations among siblings. Specifically, we calculated Spearman correlations between an individual’s NAC-shell-MS and the mean corresponding scores of their siblings across 53 families (mean family size = 2.4 ± 0.63 siblings), including 54 individuals with 22q11DS and 73 unaffected siblings. Parallel correlations were computed for BMI-Trajectory Scores (32 families, mean = 2.5 ± 0.72) and CPT-Impulsivity-MS (51 families, mean = 2.4 ± 0.63). We also computed cross-trait correlations between familial and subject-level NAC-shell-MS, BMI-Trajectory Scores, and CPT-Impulsivity-MS. Results are reported in **Supplementary Table 1** and described below.

We then conducted five additional mediation analyses to explore pathways linking familial and subject-level NAC-shell-MS and BMI-Trajectory Scores. We first tested whether the effects of Familial-CPT-Impulsivity-MS **(Model-4**), Familial-NAC-shell-MS **(Model-5)**, and Familial-BMI-Trajectory Scores **(Model-6)** on an individual’s Self-BMI-Trajectory Score were mediated by Self-NAC-shell-MS. Given significant associations observed between Familial-BMI-Trajectory Scores and Familial-NAC-shell-MS, we additionally tested whether (i) the association between Self-NAC-shell-MS and Self-BMI-Trajectory Score was mediated by Familial-NAC-shell-MS **(Model-7)**, and (ii) whether the association between Familial-NAC-shell-MS and Self-BMI-Trajectory Score was mediated by Familial-BMI-Trajectory Scores **(Model-8)**. Results are reported in **Supplementary Table 2** and described below.

#### 6.2 Results

Results of **Model 1** confirmed that NAC-shell-MS significantly mediated the association between average CPT-Impulsivity-MS and longitudinal BMI-Trajectory Scores (β = 0.06, 95% CI = [0.02–0.10], p = 0.02; Supplementary Table 2, Path 1).

Results of **Model 2** revealed that vulnerability to atypical NAC-shell maturation was a significant partial mediator of the 22q11DS effect on CPT-Impulsivity-MS (β = 0.05, 95% CI = [0.02–0.10], p = 0.002; Supplementary Table 2, Path 2).

Results of **Model 3** indicated that vulnerability to atypical NAC-shell maturation completely mediated the effect of 22q11DS diagnosis on longitudinal BMI-Trajectory Scores (β = 0.05, 95% CI = [0.01–0.11], p = 0.008; Supplementary Table 2, Path 3).

When exploring the influence of familial genetic factors beyond the 22q11.2 locus, significant familial correlations emerged between Self and Family NAC-shell-MS (ρ = 0.25, p = 0.004, Supplementary-Figure-5-Panel-A2), Self and Family BMI-Trajectory Scores (ρ = 0.36, p = 0.001, Supplementary-Figure-5-Panel-A3), and Self and Family CPT-Impulsivity-MS (ρ = 0.2, p = 0.03, Supplementary-Figure-5-Panel-A1). Additional cross-trait associations were also observed—between Family BMI-Trajectory Scores and Family-NAC-shell-MS, Family-NAC-shell-MS and Self-BMI-Trajectory, and Family-CPT-Impulsivity-MS and Self-NAC-shell-MS. A detailed description of these cross-trait associations is provided in **Supplementary Table 1**. These results support shared familial influences on NAC-shell maturation, impulsivity, and BMI trajectories.

We next conducted mediation analyses to test whether familial effects on BMI and impulsivity operated through NAC-shell maturation.

Results of **Model 4** revealed that Self-NAC-shell-MS mediated the effects of Familial-NAC-shell-MS on Self-BMI-Trajectory Score (β = –0.08, 95% CI = [–0.17 to –0.02], p = 0.06; Supplementary Table 2, Path 4).

Results of **Model 5** showed that Self-NAC-shell-MS also mediated the effects of Familial-CPT-Impulsivity-MS on Self-BMI-Trajectory Score (β = –0.08, 95% CI = [–0.18 to –0.01], p = 0.002; Supplementary Table 2, Path 5).

Results of **Model 6 instead** indicated that NAC-Shell-MS and Familial-BMI-Trajectory Scores exerted independent effects on Self-BMI-Trajectory Scores, both remaining significant when accounting for each other (Supplementary Table 2, Path 6).

Results of **Model 7** demonstrated that Familial-NAC-shell-MS mediated the association between Self-NAC-shell-MS and Family-BMI (β = –0.10, 95% CI = [–1.89 to –0.02], p = 0.002; Supplementary Table 2, Path 7).

Results of **Model 8** indicated that Family-BMI contributed to mediating the effects of Family-NAC on Self-BMI (β = –0.14, 95% CI = [–0.29 to –0.03], p = 0.002; Supplementary Table 2, Path 8).

Overall, results from Models 4, 5, and 7 indicate that NAC-shell maturation is influenced by heritable factors shared among siblings and helps explain the familial co-occurrence of impulsivity and atypical BMI. Results from Models 6 and 8 suggest the existence of partially independent mechanisms contributing to familial correlations in BMI trajectories, which remain significant after accounting for NAC-Shell-MS. These findings motivated further analyses, described in the main text, to characterize the combined impact of Familial-BMI-Trajectory and Self-NAC-Shell-MS on Self-BMI-Trajectory Scores.

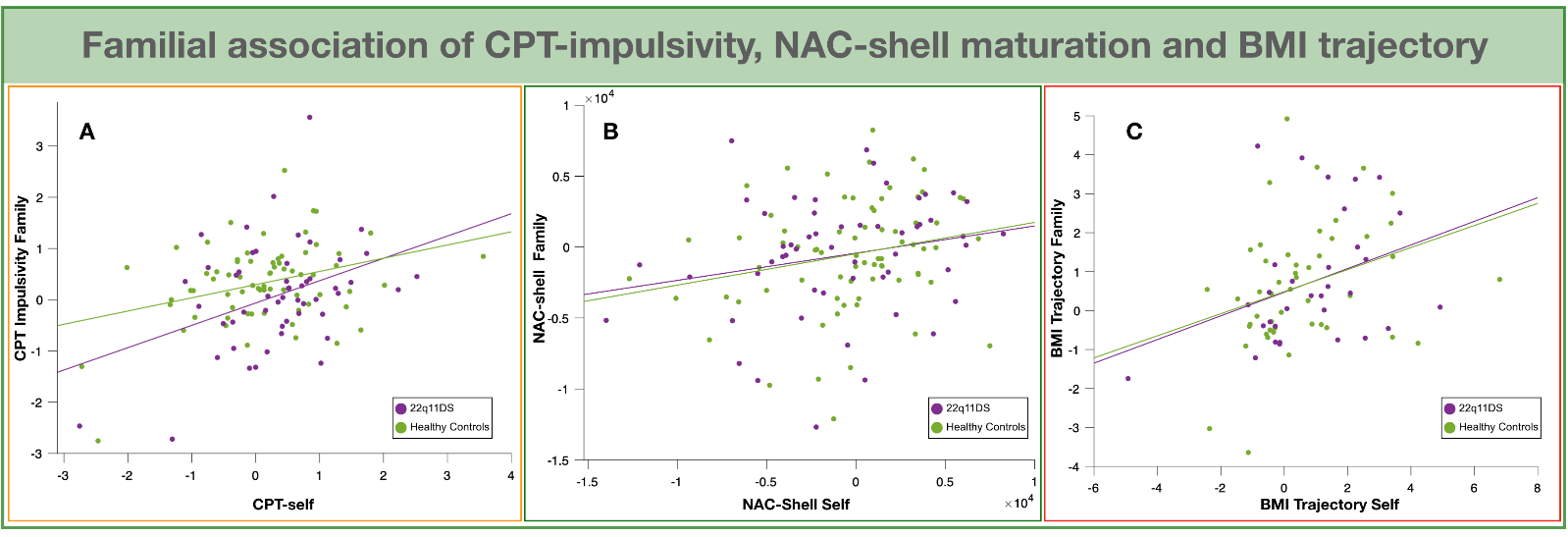

Supplementary Figure 5: Familial association of CPT-impulsivity, NAC-shell maturation and BMI trajectory

**Panel A**: Correlation between Familial CPT-Impulsivity-MS and subject-level CPT-Impulsivity-MS. **Panel B**: Correlation between Familial NAC-Shell-MS and subject-level NAC-Shell-MS. **Panel C**: Correlation between Familial BMI trajectory and subject-level BMI trajectory.

|  | NAC self | | CPT self | | BMI self | | NAC family | | CPT family | | BMI family | |
| --- | --- | --- | --- | --- | --- | --- | --- | --- | --- | --- | --- | --- |
|  | rho | p-value | rho | p-value | rho | p-value | rho | p-value | rho | p-value | rho | p-value |
| NAC self |  |  | **-0.26** | **<0.001** | **-0.36** | **<0.001** | **0.25** | **0.004** | **-0.24** | **0.009** | -0.12 | 0.28 |
| CPT self | **-0.26** | **<0.001** |  |  | 0.07 | 0.33 | **-0.24** | **0.008** | 0.2 | **0.029** | 0.07 | 0.55 |
| BMI self | **-0.36** | **<0.001** | 0.07 | 0.33 |  |  | -0.1 | 0.31 | -0.003 | 0.98 | **0.36** | **0.001** |
| NAC family | **0.25** | **0.004** | **-0.24** | **0.008** | -0.11 | 0.1 |  |  | -0.16 | 0.08 | **-0.46** | **<0.001** |
| CPT family | **-0.24** | **0.009** | **0.2** | **0.029** | -0.003 | 0.98 | -0.16 | 0.08 |  |  | -0.11 | 0.34 |
| BMI family | -0.12 | 0.28 | 0.07 | 0.55 | **0.36** | **0.001** | **-0.46** | **<0.001** | -0.11 | 0.34 |  |  |

Supplementary Table 1 : Homologous and cross-trait associations between familial and subject-level self NAC-Shell-MS, BMI-Trajectory-Scores and CPT-Impulsivity-MS.

Values represent Spearman’s rho coefficients and the associated p-value. Significant correlations are shown in bold.

| **Path-1: 3-way Mediation Analysis of Self CPT impulsivity, Self NAC-shell maturation and Self BMI trajectory** | | | | |
| --- | --- | --- | --- | --- |
|  | *Estimate* | *p-value* | *Lower CI* | *Upper CI* |
| *CPT impulsivity self (path a)* | **-0.19** | **0.01** | **-0.34** | **-0.46** |
| *CPT impulsivity self (path c’)* | 0.03 | 0.69 | -0.11 | 0.17 |
| *NAC-shell self (path b)* | **-0.29** | **0.0001** | **-0.43** | **-0.14** |
| *CPT impulsivity self (path c)* | 0.08 | 0.26 | -0.063 | 0.23 |
| *a*b* | **0.055** | **0.016** | **0.015** | **0.1** |
| **Path 2: 3-way Mediation Analysis of 22q11DS diagnosis, Self NAC-shell maturation and Self CPT impulsivity** | | | | |
|  | *Estimate* | *p-value* | *Lower CI* | *Upper CI* |
| *22q11DS diagnosis (path a)* | **-0.24** | **0.0001** | **-0.36** | **-0.12** |
| *22q11DS diagnosis (path c’)* | 0.078 | 0.22 | -0.047 | 0.2 |
| *NAC-shell self (path b)* | **-0.21** | **0.0008** | **-0.34** | **-0.09** |
| *22q11DS diagnosis (path c)* | **0.13** | **0.04** | **0.006** | **0.25** |
| *a*b* | **0.05** | **0.002** | **0.02** | **0.1** |
| **Path-3: 3-way Mediation Analysis of 22q11DS diagnosis, Self NAC-shell maturation and Self BMI trajectory** | | | | |
|  | *Estimate* | *p-value* | *Lower CI* | *Upper CI* |
| *22q11DS diagnosis (path a)* | **-0.19** | **0.01** | **-0.34** | **-0.05** |
| *22q11DS diagnosis (path c’)* | 0.08 | 0.27 | -0.06 | 0.22 |
| *NAC-shell self (path b)* | **-0.28** | **0.0002** | **-0.42** | **-0.13** |
| *22q11DS diagnosis (path c)* | 0.13 | 0.07 | -0.013 | 0.28 |
| *a*b* | **0.05** | **0.008** | **0.01** | **0.11** |
| **Path-4: 3-way Mediation Analysis of Familial NAC-shell maturation, Self NAC-shell maturation and Self BMI trajectory** | | | | |
|  | *Estimate* | *p-value* | *Lower CI* | *Upper CI* |
| *NAC-shell Family (path a)* | **0.22** | **0.01** | **0.05** | **0.4** |
| *NAC-shell Family (path c’)* | -0.036 | 0.72 | -0.24 | 0.16 |
| *NAC-shell self (path b)* | **-0.37** | **0.0004** | **-0.56** | **-0.17** |
| *NAC-shell Family (path c)* | -0.11 | 0.29 | -0.32 | 0.1 |
| *a*b* | **-0.08** | **0.006** | **-0.17** | **-0.02** |
| **Path-5: 3-way Mediation Analysis of Familial CPT impulsivity, Self NAC-shell maturation and Self BMI trajectory** | | | | |
|  | *Estimate* | *p-value* | *Lower CI* | *Upper CI* |
| *CPT impulsivity Family (path a)* | **-0.21** | **0.02** | **-0.39** | **-0.036** |
| *CPT impulsivity Family (path c’)* | -0.009 | 0.92 | -0.2 | 0.19 |
| *NAC-shell self (path b)* | **-0.37** | **0.0004** | **-0.56** | **-0.17** |
| *CPT impulsivity Family (path c)* | 0.035 | 0.7 | -0.17 | 0.24 |
| *a*b* | **0.08** | **0.018** | **0.01** | **0.18** |
| Path-6: 3-way Mediation Analysis of Familial BMI trajectory, Self NAC-shell maturation and Self BMI trajectory | | | | |
|  | Estimate | p-value | Lower CI | Upper CI |
| *BMI trajectory Family (path a)* | -0.14 | 0.21 | -0.37 | 0.08 |
| *BMI trajectory Family (path c’)* | **0.29** | **0.007** | **0.08** | **0.49** |
| *NAC-shell self (path b)* | **-0.32** | **0.002** | **-0.52** | **-0.12** |
| *BMI trajectory Family (path c)* | **0.33** | **0.003** | **0.12** | **0.55** |
| *a*b* | 0.049 | 0.1 | -0.019 | 0.14 |
| **Path-7: 3-way Mediation Analysis of Self NAC-shell maturation, Familial NAC-shell maturation and Familial BMI trajectory** | | | | |
|  | Estimate | p-value | Lower CI | Upper CI |
| *NAC-shell self (path a)* | **0.22** | **0.01** | **0.05** | **0.39** |
| *NAC-shell self (path c’)* | -0.07 | 0.5 | -0.27 | 0.14 |
| *NAC-shell Family (path b)* | **-0.43** | **0.0001** | **-0.64** | **-0.22** |
| *NAC-shell self (path c)* | -0.14 | 0.2 | -0.36 | 0.08 |
| *a*b* | **-0.1** | **0.002** | **-1.89** | **-0.02** |
| **Path-8: 3-way Mediation Analysis of Familial NAC-shell maturation, Familial BMI trajectory and Self BMI trajectory** | | | | |
|  | *Estimate* | *p-value* | *Lower CI* | *Upper CI* |
| *NAC-shell Family (path a)* | **-0.44** | **6.36*10^-5^** | **-0.64** | **-0.23** |
| *NAC-shell Family (path c’)* | -0.03 | 0.8 | -0.27 | 0.21 |
| *BMI trajectory Family (path b)* | **0.32** | **0.009** | **0.08** | **0.56** |
| *NAC-shell Family (path c)* | -0.1 | 0.29 | -0.32 | 0.1 |
| *a*b* | **-0.14** | **0.002** | **-0.29** | **-0.03** |

Supplementary Table 2: Detailed statistics for all mediation analyses reported in the maintext.
